# Relationship between Retinal Vessels and OCT-Derived Retinal Neural Parameters

**DOI:** 10.1101/2024.09.23.24313858

**Authors:** Mayinuer Yusufu, Robert N. Weinreb, Mengtian Kang, Algis J. Vingrys, Xianwen Shang, Lei Zhang, Danli Shi, Mingguang He

## Abstract

**Objective:** To investigate structural relationships between retinal vasculometry derived from color fundus photography (CFP) and neural parameters obtained from Optical Coherence Tomography (OCT) scans and validate their causal relationships.

**Design:** Cross-sectional study

**Participants:** Participants with fundus photographs data and OCT data in the UK Biobank cohort study

**Methods:** We used the Retina-based Microvascular Health Assessment System (RMHAS) to extract retinal vascular measurements in the 6^*^6mm area centered on the macular region. OCT parameters were available from the UK Biobank. First, pairwise correlations between individual retinal layers and vascular parameters were investigated. Canonical correlation analysis (CCA) was then used to examine associations between sets of variables. Lastly, bidirectional two-sample Mendelian randomization was employed to investigate potential causal relationships.

**Main Outcome Measures:** Measurements of retinal vascular network and neural layers

**Results:** Data from 67,918 eyes of 43,029 participants were included. The Ganglion Cell-Inner Plexiform Layer (GC-IPL) thickness showed the strongest correlations with vascular Density and Complexity (r=0.199 for arterial Vessel Area Density and r=0.175 for Number of Segments). The Inner Nuclear Layer (INL) thickness showed positive correlations with Width (r=0.122) and Vessel Area Density (artery) (r=0.127). Mendelian randomization analysis indicated bidirectional causal relationships between retinal vascular features and layer thicknesses. Genetically predicted higher Vessel Density was associated with increased thickness across various retinal layers, with the strongest effect on Inner Segment/Outer Segment + Photoreceptor Segment thickness (standardized effect size 1.50, p<0.001). Genetically predicted increases in retinal layer thicknesses, particularly the Outer Plexiform Layer, were linked to higher Vessel Density (standardized effect size 0.45, p=0.002) and Fractal Dimension (standardized effect size 0.48, p<0.001).

**Conclusions:** The positive associations of macular thickness with vascular Density and Caliber measurements were mainly attributable to their associations with GC-IPL and INL. Multidimensional relationships revealed by CCA revealed a complementary nature between the two sets of parameters, highlighting their value as a composite biomarker. Mendelian Randomization uncovered a bidirectional causal relationship that should provide insights into novel therapeutic approaches targeting both vascular and neuronal components.

## Introduction

The retina, an extension of the central nervous system, offers a unique window into human physiology. It allows for direct, non-invasive visualization of its vasculature and neural structures^1,2^, lending itself as a valuable proxy for assessing both ocular and systemic health^3-5^. For instance, retinal vascular features, including vessel diameter, tortuosity, and branching patterns, quantified from color fundus photography (CFP), have been associated with cardiovascular diseases, diabetes, and hypertension ^6,7^. Retinal layer measurements such as retinal nerve fiber layer (RNFL) thickness and ganglion cell-inner plexiform layer (GC-IPL) thickness derived from Optical Coherence Tomography (OCT) have shown associations with neurodegenerative diseases like Alzheimer’s disease, Parkinson’s disease, and multiple sclerosis^8-10^.

While previous studies have explored associations between retinal features and various systemic conditions, they primarily focused on studying retinal vascular and neural features separately. There is a gap in our understanding of the relationships between the information provided by those two modalities. Although the development of artificial intelligence has empowered their detailed quantification and facilitated the differentiation of subtle retinal feature alterations,^11,12^ there is a paucity of information about detailed associations between vascular features observed in CFP and neural characteristics measured by OCT.

CFP and OCT offer complementary data on vascular and neural health, respectively. Notably, the functional interplay between retinal vessels and neurons and ganglion cells, known as neurovascular coupling, plays an essential role in maintaining normal retinal physiology and is disrupted in many ocular diseases, such as diabetic retinopathy and glaucoma^13,14^, and by extension, brain function^15^. In addition, their interrelationship could provide a more comprehensive understanding of retinal physiology and pathology, as well as the underlying pathophysiology of conditions involving both vascular and neural health. Such insights could lead to the identification of early composite markers of neurodegeneration and vascular cognitive impairment, facilitating earlier diagnosis and intervention in conditions such as Alzheimer’s disease^16,17^. Furthermore, it remains unclear which process (vascular or neural structure changes) precedes the other. One hypothesis posits that nerve damage leads to reduced energy usage, subsequently causing a decrease in vessel density to match the lowered metabolic demand^18^. Conversely, it is possible that vascular damage could precede and contribute to nerve degeneration by compromising blood supply and nutrient delivery to neural tissues ^19^.

In the current study, we aim to bridge this gap by (1) investigating structural relationships between retinal vasculometry, obtained from CFPs using the Retina-based Microvascular Health Assessment System (RMHAS)^11^, and OCT-derived retinal layer parameters in a large cohort of participants from the UK Biobank; (2) Unveil the causal relationship between retinal vascular and neural feature changes through Mendelian Randomization.

## Methods

### Study design and population

For the correlation analyses in this cross-sectional study, we used unidentifiable data from the UK Biobank. The UK Biobank study is a large population-based cohort study that enrolled participants aged 40 to 69 years that was launched in 2006 in the United Kingdom, and introduced eye examinations in 2009, including CFP^20,21^. The UK Biobank cohort study obtained ethical approval from the North West Multi-Centre Research Ethics Committee (reference number 06/MRE08/65).

### Inclusion and exclusion criteria

We excluded participants who withdrew their consent or did not have CFPs and/or OCT data. The quality of the CFPs was assessed with RMHAS, and those classified as “Reject” were removed^11^. The quality of OCT data was assessed using the quality control indicators obtained with the Topcon Advanced Boundary Segmentation software^22^. Images with a quality score less than 45, within the poorest 20% of centration and segmentation uncertainty were excluded. Lastly, we excluded eyes with conditions that could potentially affect vascular network quantification or retinal layer thickness measurements. These included intraocular pressure (IOP) of ≥21 mmHg or ≤8 mmHg, self-reported glaucoma, macular degeneration, retinal diseases, diabetic eye diseases, or history of glaucoma surgery^12^. (Definitions are presented in Supplementary Table 1)

### Retinal vascular Measurements

The retinal vascular parameters were obtained using RMHAS^11^ from a 6^*^6mm area centered on the macular region of CFPs. The RMHAS automatedly segmented and quantified 23 measurements of 5 categories: Calibers, Density, Tortuosity, Branching Angle, and Complexity. Supplementary Table 2 shows the definition of retinal vascular features. Figure 1 shows the corresponding structure of the retinal vascular network obtained from CFPs and cross-sectional layers obtained from OCT scans in the 6^*^6mm area.

**Figure 1.**
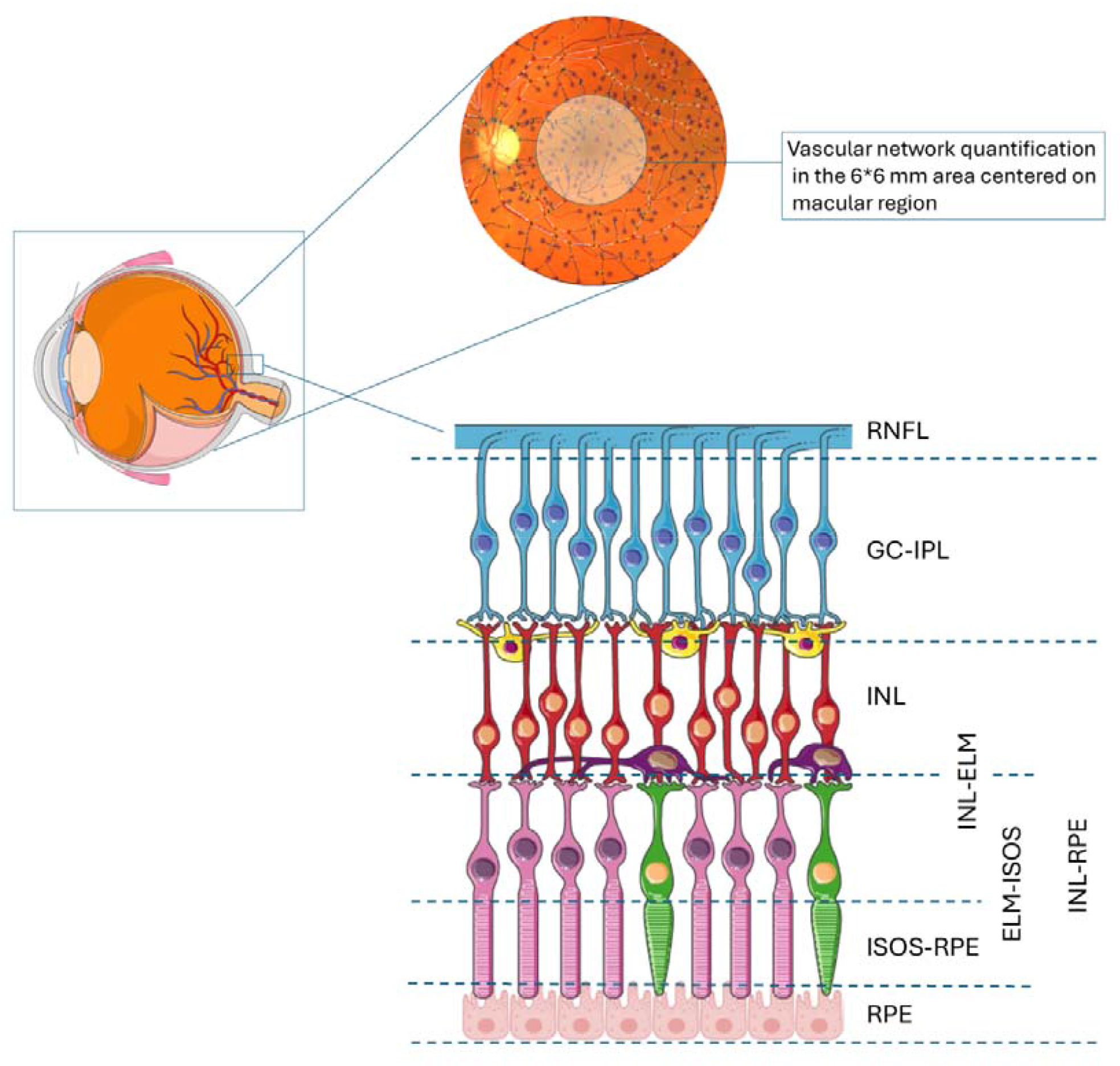
Two imaging modalities of retina structures Notes: The figure used images downloaded from the Servier Medical Art https://smart.servier.com/smart_image and by Sally Kim. (Structure of the Retina.: Biorender; 2020. https://app.biorender.com/biorender-templates/figures/all/t-5fdba689c542b300a3aeb236-structure-of-the-retina). Abbreviations: RNFL, retinal nerve fiber layer; GC-IPL, ganglion cell layer-inner plexiform layer; INL, inner nuclear layer; ELM, external limiting membrane; ISOS, inner segment/outer segment; RPE, retinal pigment epithelium.

**Figure 2.**
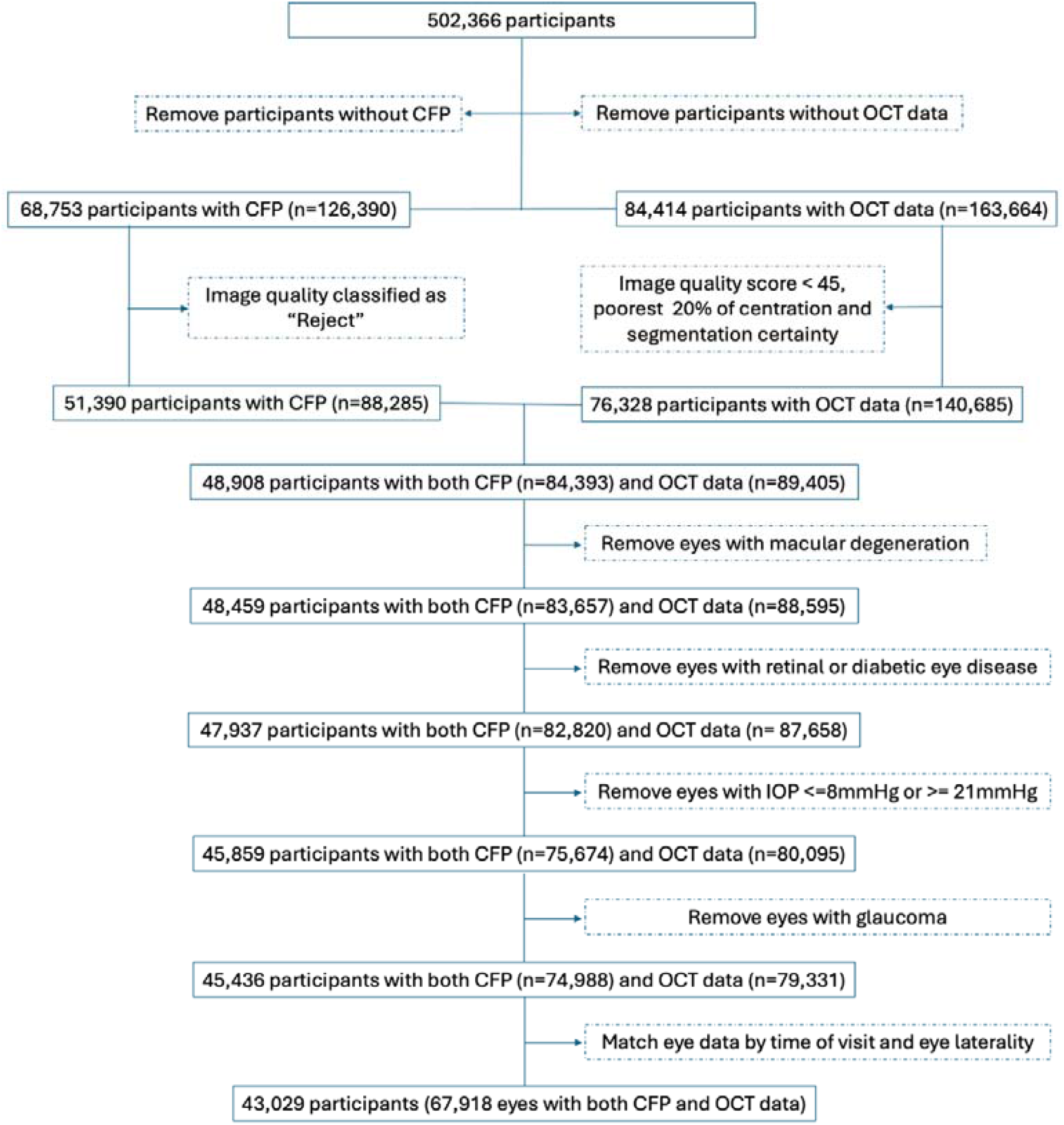
Participant selection process Notes: OCT, Optical Coherence Tomography. CFP, Color Fundus Photograph. Definitions of variables used in the selection process are presented in Supplementary Table 1.

### OCT Parameters

OCT images were acquired using spectral domain OCT (Topcon 3D OCT 1000 Mk2, Topcon Inc, Oakland, NJ, USA), capturing high-resolution, cross-sectional scans of the retina, 6 mm x 6 mm area centered on the fovea, without pupillary dilation in a dark room setting^23^. OCT parameters were extracted using Topcon Advanced Boundary Segmentation software^12^. After removing parameters with more than 30% missing values and keeping only parameters presenting retinal layer thickness of the 6^*^6mm area, we included 9 parameters: macular thickness, RNFL, GC-IPL, inner nuclear layer (INL), INL-retinal pigment epithelium (RPE), INL-external limiting membrane (ELM), ELM-inner segment/outer segment (ISOS), ISOS-RPE, and RPE thickness (Figure 1).

### Mendelian Randomization

We performed a bidirectional, two-sample Mendelian Randomization to examine further the potential causal relationship between vascular network features and retinal layer thickness. For the vascular features, we used single nucleotide polymorphisms (SNPs) of retinal vascular network features obtained in the UK Biobank^24^. The study^24^ provided genome-wide association study (GWAS) data on retinal Vessel Density and Fractal Dimension. For the retinal layer thickness, we used GWAS data of retinal layer thickness obtained from the Leipzig Research Centre for Civilization Diseases (LIFE-Adult Study)^25,26^. Details of GWAS studies can be found in Supplementary Text 1. The baseline of the LIFE-Adult Study was carried out from August 2011 to November 2014 in Leipzig, Germany, focusing on investigating prevalences, early onset markers, genetic predispositions, and lifestyle determinants of major civilization diseases in participants aged 40-79^26^.

### Statistical analysis

To describe baseline characteristics, we summarized continuous variables with mean (standard deviation [SD]), while categorical variables were presented as counts and percentages. We removed outliers using the method proposed by Zekayat et al^24^. Parameters with a missing proportion of > 30% were excluded. Missing values were imputed using Multivariate Imputation by Chained Equations. To ensure comparability, all measurements were rescaled to the SD unit.

We assessed normality of the data distribution was assessed with the Anderson-Darling test. For the correlation test, we used Pearson correlation if both variables were normally distributed, otherwise Spearman’s rank correlation was used. We generated a heatmap based on the correlation matrix to assess the pairwise strength and direction of the relationships between all possible pairs of retinal vessel characteristics and OCT parameters.

For broader structural relationships between these two sets of variables, we employed Canonical Correlation Analysis (CCA). The CCA generates dimensions that are linear combinations of variables from each set that maximize the correlation between the two sets. The proportion of variance explained by each canonical dimension was calculated to assess the relative importance of each dimension in explaining the overall relationship between the two sets of variables. The scatter plot with a fitted locally estimated scatterplot smoothing (LOESS) curve and linear regression line illustrated the correlation in the canonical dimensions. A Sankey diagram was constructed to depict the change in variable importance across canonical dimensions.

For the bidirectional Mendelian Randomization, SNPs with a GWAS-correlated P-value < 5×10^−6^ were selected, and data clumping was performed with the linkage disequilibrium r^2^ set at 0.001 and clumping window set at 10000 kb. To ensure the comparability among parameters, the standardized effect sizes were presented.

Additionally, subgroup analyses by sex, age groups, and eye laterality were performed. In this study, a two-sided significance level of alpha = 0.05 was set for all statistical tests. All analyses were conducted using R version 4.2.3.

## Results

### Characteristics of participants

Starting with 502,366 participants, our selection process filtered for those with both CFP and OCT data but excluded low-quality images and data and eyes with conditions that would affect the segmentation and quantification of retinal features. Our final cohort was 43,029 participants with 67,918 eyes that had both quality CFP and OCT data.

The mean (SD) age of the participants was 55.5 (8.19) years, with a higher proportion of females (55.6%) than males (44.4%) and males being slightly older than females (55.7 vs 55.3 years, p<0.001). Males had a higher mean body mass index (27.6 vs 26.7 kg/m^2^, p<0.001) and were more likely to be current or former smokers (47.1% vs 38.2%, p<0.001) compared with females. Males also reported higher levels of physical activity, with 36.0% in the high category compared with 30.6% of females (p<0.001). Cardiovascular risk factors showed significant differences across genders, with males having higher mean SBP (138 vs 133 mmHg, p<0.001), DBP (83.2 vs 79.8 mmHg, p<0.001), and HbA1c levels (35.8 vs 35.3 mmol/mol, p<0.001). Females had higher mean high-density lipoprotein (1.63 vs 1.31 mmol/L, p<0.001) and low-density lipoprotein (3.59 vs 3.51 mmol/L, p<0.001) levels. The prevalence of diabetes (4.9% vs 2.8%, p<0.001) and cardiovascular diseases (29.9% vs 21.3%, p<0.001) was higher in males. Detailed characteristics of participants stratified by sex can be found in Table 1.

**Table 1.** Baseline characteristics of participants.

| Characteristic | Whole Sample<br>(N=43029) | Female<br>(N=23917) | Male<br>(N=19112) | P-value |
| --- | --- | --- | --- | --- |
| **Age (years)** |  |  |  | <0.001 |
| Mean (SD) | 55.5 (8.19) | 55.3 (8.02) | 55.7 (8.38) |  |
| **Body Mass Index**<br>**(kg/m<sup>2</sup>)** |  |  |  | <0.001 |
| Mean (SD) | 27.1 (4.69) | 26.7 (5.09) | 27.6 (4.10) |  |
| Missing, N (%) | 202 (0.5%) | 107 (0.4%) | 95 (0.5%) |  |
| **Smoking Status, N (%)** |  |  |  | <0.001 |
| Never | 24664 (57.3%) | 14665 (61.3%) | 9999 (52.3%) |  |
| Previous | 14191 (33.0%) | 7264 (30.4%) | 6927 (36.2%) |  |
| Current | 3940 (9.2%) | 1862 (7.8%) | 2078 (10.9%) |  |
| Missing | 234 (0.5%) | 126 (0.5%) | 108 (0.6%) |  |
| **Physical Activity, N (%)** |  |  |  | <0.001 |
| Low | 6107 (14.2%) | 3184 (13.3%) | 2923 (15.3%) |  |
| Moderate | 14124 (32.8%) | 8027 (33.6%) | 6097 (31.9%) |  |
| High | 14199 (33.0%) | 7310 (30.6%) | 6889 (36.0%) |  |
| Missing | 8599 (20.0%) | 5396 (22.6%) | 3203 (16.8%) |  |
| **Systolic Blood Pressure**<br>**(mmHg)** |  |  |  | <0.001 |
| Mean (SD) | 135 (18.0) | 133 (18.6) | 138 (16.6) |  |
| Missing, N (%) | 144 (0.3%) | 90 (0.4%) | 54 (0.3%) |  |
| **Diastolic Blood Pressure**<br>**(mmHg)** |  |  |  | <0.001 |
| Mean (SD) | 81.3 (9.98) | 79.8 (9.92) | 83.2 (9.74) |  |
| Missing, N (%) | 144 (0.3%) | 90 (0.4%) | 54 (0.3%) |  |
| **Glycated Haemoglobin**<br>**(mmol/mol)** |  |  |  | <0.001 |
| Mean (SD) | 35.6 (5.79) | 35.3 (5.19) | 35.8 (6.44) |  |
| Missing, N (%) | 5127 (11.9%) | 2938 (12.3%) | 2189 (11.5%) |  |
| **High-Density**<br>**Lipoprotein (mmol/L)** |  |  |  | <0.001 |
| Mean (SD) | 1.49 (0.387) | 1.63 (0.382) | 1.31 (0.312) |  |
| Missing, N (%) | 5742 (13.3%) | 3383 (14.1%) | 2359 (12.3%) |  |
| **Low-Density Lipoprotein**<br>**(mmol/L)** |  |  |  | <0.001 |
| Mean (SD) | 3.55 (0.852) | 3.59 (0.853) | 3.51 (0.849) |  |
| Missing, N (%) | 3677 (8.5%) | 2117 (8.9%) | 1560 (8.2%) |  |
| **Diabetes, N (%)** |  |  |  | <0.001 |
| No | 41205 (95.8%) | 23141 (96.8%) | 18064<br>(94.5%) |  |
| Yes | 1589 (3.7%) | 662 (2.8%) | 927 (4.9%) |  |
| Missing | 235 (0.5%) | 114 (0.5%) | 121 (0.6%) |  |
| **CVD, N (%)** |  |  |  | <0.001 |
| No | 32024 (74.4%) | 18709 (78.2%) | 13315<br>(69.7%) |  |
| Yes | 10813 (25.1%) | 5097 (21.3%) | 5716 (29.9%) |  |
| Missing | 192 (0.4%) | 111 (0.5%) | 81 (0.4%) |  |

Notes: SD: standard deviation; N, number; CVD, Cardiovascular Disease

### Pairwise correlations

Figure 3 illustrates the pairwise correlations between retinal vascular features and the thickness of retinal layers. Macular thickness had moderate positive correlations with multiple vascular features, particularly the Density measure. The strongest correlations were observed for Vessel Area Density (artery) (r=0.161), Vessel Skeleton Density (artery) (r=0.132), and Width (r=0.118). Similarly, GC-IPL demonstrated even stronger positive correlations with these vascular features, with the highest correlations observed for Vessel Area Density (artery) (r=0.199), Vessel Skeleton Density (artery) (r=0.170), and Vessel Skeleton Density (vein) (r=0.152). INL also showed moderate positive correlations with several vascular features, such as Width (r=0.122) and Vessel Area Density (artery) (r=0.127).

**Figure 3.**
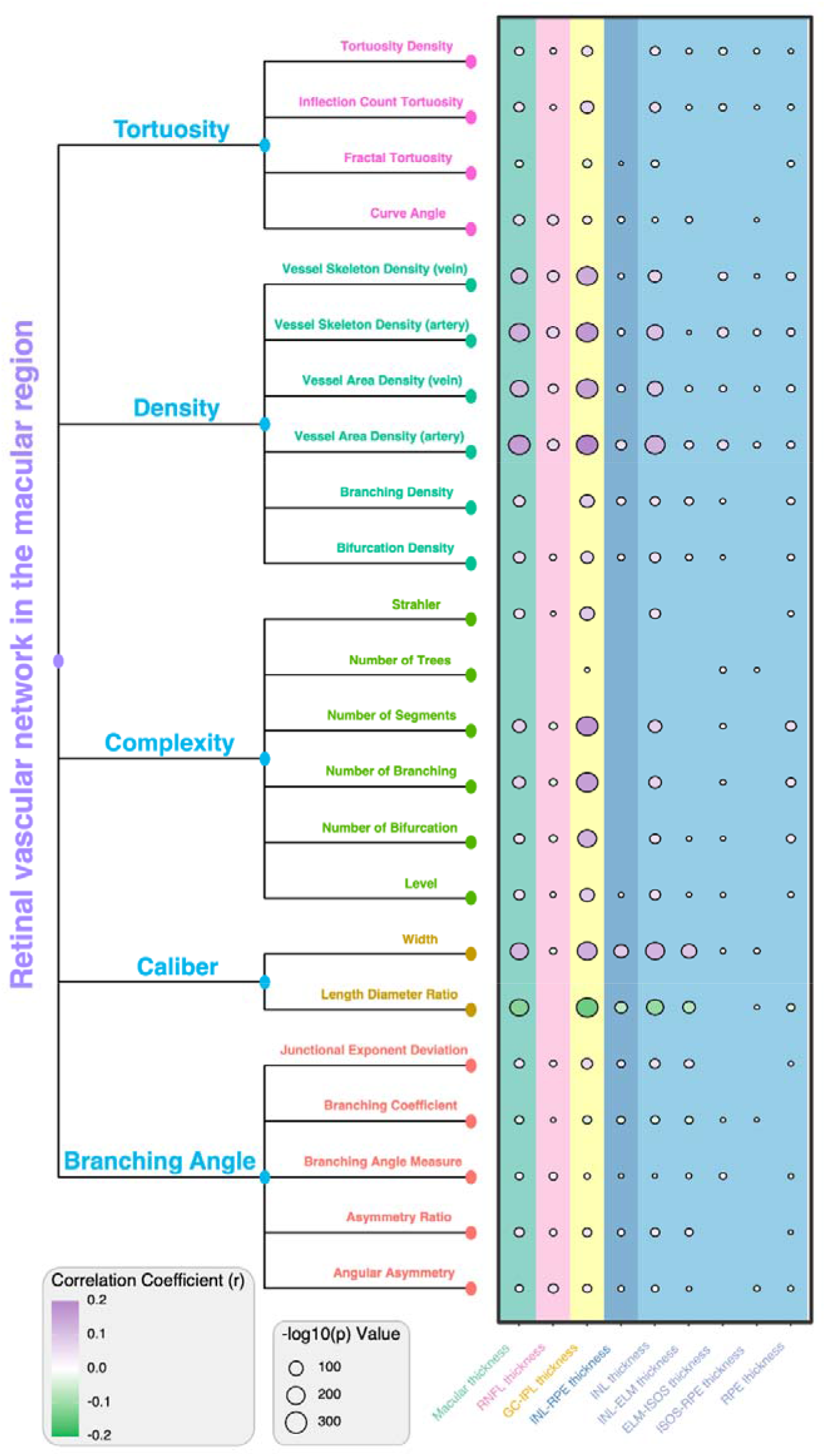
Correlations between retinal vascular features and thickness of retinal layers Notes: Circle size represents statistical significance (-log10(p) value), while its color indicates the strength and direction of correlation (purple for positive, green for negative). Only those correlations with p<0.05 were presented with a circle. Abbreviations: RNFL, retinal nerve fiber layer; GC-IPL, ganglion cell-inner plexiform layer; INL, inner nuclear layer; ELM, external limiting membrane; ISOS, inner segment/outer segment; RPE, retinal pigment epithelium. The calculated correlation coefficients and the corresponding p-values can be found in Supplementary Table 3.

GC-IPL showed strong correlations with Complexity measures, such as Number of Segments (r=0.175) and Number of Branching (r=0.157). Conversely, the Length Diameter Ratio (LDR) showed negative correlations with multiple retinal layer thicknesses, particularly GC-IPL (r=−0.143) and INL (r=−0.106). RNFL shows weaker correlations, with the strongest being a modest positive correlation with Vessel Skeleton Density (artery) (r=0.065). Most correlations were statistically significant (p<0.001), although the strength of these correlations varied, with most falling in the weak to moderate range.

### Canonical Correlation Analysis

Figure 4 shows the main results of CCA. The sharp increase in cumulative variance observed in the first three dimensions suggests that most of the important relationships between the two sets of variables are captured. The first canonical dimension explained approximately 43.8% of the variance in the relationship between retinal vascular and OCT parameters, and subsequent dimensions contributed 29.88%. Taken together, the first three dimensions collectively account for over 92.02% of the total variance explained.

**Figure 4.**
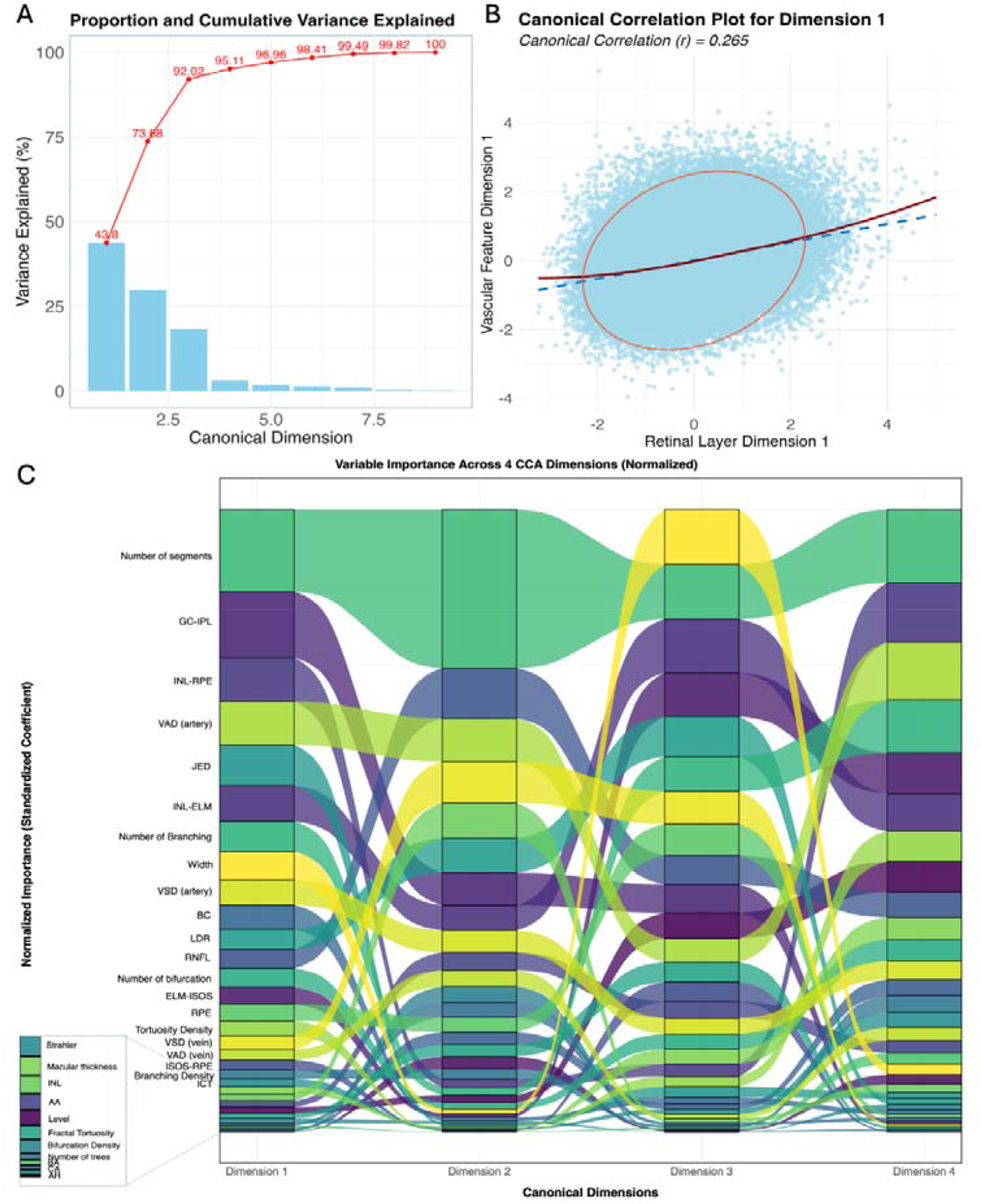
Canonical Correlation Analysis (CCA) of Retinal Vascular Measurements and Retinal Layer Thickness Notes: (A) Variance Explained: The bar chart (blue) shows the proportion of variance explained by each canonical dimension, while the line graph (red) displays the cumulative variance explained. This illustrates the relative importance of each dimension in capturing the relationship between retinal vascular and OCT parameters. The first four dimensions explained over 95% of the variance. (B) First Canonical Dimension Correlation: This scatter plot depicts the correlation between the first canonical variates for retinal vascular measurements (y-axis) and retinal layer thickness (x-axis). The red ellipse outlines the 95% confidence region of the data distribution. The blue dashed line shows the linear regression fit, while the solid red curve represents the locally estimated scatterplot smoothing (LOESS) fit, providing a non-linear visualization of the relationship. The canonical correlation (ρ) of 0.265 indicates the strength of the association for this dimension. (C) Variable Importance Across Dimensions: This Sankey diagram illustrates how the importance of different variables (both retinal vascular measurements and OCT parameters) changed across the first four canonical dimensions. The height of each flow represents the magnitude of a variable’s contribution to each dimension, allowing for the visualization of which variables are most influential in each canonical relationship. Supplementary Figures 1 and 2 show the correlations of the first four dimensions and the loadings of variables.

The first canonical dimension showed a moderate positive correlation (ρ = 0.265) between retinal vascular measurements and retinal layer thickness. The correlation coefficients were 0.218 and 0.171 in the second and third dimensions (all p <0.001). In the scatter plots of the first three canonical dimensions, the LOESS curves largely overlapped with the fitted linear lines, suggesting predominant linear relationships with merely weak non-linear relationships at the extremes of the distributions.

The Sankey diagram revealed that in the first dimension, the number of segments, GC-IPL thickness, and INL-RPE thickness appeared to be the most influential variables. The second dimension showed that Vessel Area Density (artery), Junctional Exponent Deviation (JED), and INL-ELM thickness are the major contributors. The third and fourth dimensions displayed more evenly distributed patterns of variable importance, with contributions from parameters such as vessel width, branching patterns, and various layer thicknesses.

### Mendelian Randomization

Table 2 presents the results of bidirectional Mendelian randomization, revealing significant associations of various retinal layers with Vessel Density and Fractal Dimension. Vessel Density showed the strongest effect on ISOS + Photoreceptor Segment (ISOS+PS) thickness, with a standardized effect size of 1.50 (95% CI: 1.07, 1.93; p<0.001). In addition, Vessel Density also showed significant positive effects on Ganglion Cell Layer (GCL) and Outer Plexiform Layer (OPL) thicknesses, with standardized effect sizes of 0.50 and 0.57 (both p-values <0.05). When examining the effect of layer thickness on Vessel Density, multiple retinal layer thicknesses exhibited significant effects. OPL thickness demonstrated the largest effect on Vessel Density, with an effect size of 0.45 (95% CI: 0.16, 0.75, p=0.002). GCC, GCL, ISOS+PS, and INL thicknesses also significantly positively affected Vessel Density, with effect sizes ranging from 0.27 to 0.40 (all p<0.05). While Mendelian randomization did not demonstrate the effect of Fractal Dimension on the thickness of layers, the reverse effect was revealed. GCC, GCL, IPL, OPL, and INL all showed significant effects, with standardized effect sizes ranging between 0.25 and 0.48.

**Table 2.** Bidirectional Mendelian Randomization.

| Exposure | Outcome | Method | NSNPs | Standardized Effect Size (95% CI) | P-value |
| --- | --- | --- | --- | --- | --- |
| VD | ISOS+PS | Wald ratio | 1 | 1.50 (1.07, 1.93) | <0.001 |
| VD | GCL | Wald ratio | 1 | 0.50 (0.10, 0.89) | 0.015 |
| VD | OPL | Wald ratio | 1 | 0.57 (0.37, 0.78) | <0.001 |
| OPL | VD | IVW | 3 | 0.45 (0.16, 0.75) | 0.002 |
| GCC | VD | IVW | 2 | 0.27 (0.12, 0.41) | <0.001 |
| ISOS+PS | VD | IVW | 2 | 0.28 (0.20, 0.35) | <0.001 |
| GCL | VD | IVW | 3 | 0.40 (0.06, 0.75) | 0.019 |
| INL | VD | Wald ratio | 1 | 0.39 (0.32, 0.46) | <0.001 |
| IPL | VD | IVW | 2 | 0.43 (-0.02, 0.87) | 0.06 |
| GCL | FD | IVW | 2 | 0.44 (0.29, 0.58) | <0.001 |
| INL | FD | Wald ratio | 1 | 0.25 (0.18, 0.32) | <0.001 |
| GCC | FD | Wald ratio | 1 | 0.32 (0.19, 0.46) | <0.001 |
| IPL | FD | Wald ratio | 1 | 0.44 (0.32, 0.56) | <0.001 |
| OPL | FD | Wald ratio | 1 | 0.48 (0.35, 0.61) | <0.001 |

Notes: SNP, single nucleotide polymorphism; NSNPs, number of SNPs; VD, Vessel Density; FD, Fractal Dimension; OPL, outer plexiform layer; GCC, ganglion cell complex; ISOS, inner segment/outer segment; PS photoreceptor segments; GCL, ganglion cell layer; INL, inner nuclear layer; GCC included RNFL, GCL, and inner plexiform layer. ISOS+PS included external limiting membrane-ISOS + ISOS-retinal pigment epithelium; IVW, Inverse variance weighted. When only one SNP was identified for each pair of exposure and outcome, the Wald ratio was used, while if more than one was identified, then IVW was used. The standardized effect size was calculated to represent the change in the outcome variable (in SD units) for a one-unit change in the exposure variable.

## Discussion

We conducted a comprehensive analysis of the relationships between retinal layer thicknesses and vascular features in 67,918 eyes of 43,029 participants by evaluating pairwise correlations, correlations between two sets of parameters, and their genetic influence on each other. The GC-IPL and INL had the strongest associations with vascular features, including Density, Complexity, and Caliber measurements; there were stronger correlations with arteries than veins. The CCA results further revealed multidimensional relationships between the two sets of parameters, indicating a complementary nature of their relationships. Furthermore, the Mendelian Randomization analysis provided evidence for positive bidirectional causal relationships.

The pairwise correlation analysis revealed the correlations between macular thickness (mainly attributable to GC-IPL and INL thickness) and Density, Caliber, and Complexity measurements. The strongest correlations were found for GC-IPL with Vessel Area Density (artery) and Vessel Skeleton Density (artery), Number of Segments, and Number of Branching. The Mendelian Randomization also revealed the effect of Vessel Density on GCL. This suggests that a richer blood supply in the macular region could support enhanced growth of ganglion cells. Given the high metabolic demand of ganglion cell bodies and dendrites in the GC-IPL, its thickness is critically dependent on adequate blood supply^19,27,28^. This also explains the stronger correlation found for arterial Density parameters compared with venular Density parameters in our study. Furthermore, as revealed by the bidirectional Mendelian Randomization, genetically determined thicker ganglion cell complex can result in higher Vessel Density and vice versa. These findings align with previous research suggesting the intricate bidirectional interaction between retinal vascular health and blood supply regulation is closely linked to the integrity of retinal neurons, and neural acticity^18,29-31^, providing additional evidence on neurovascular coupling in the retina in terms of phenotypic morphology and genetics.

INL thickness was another layer primarily contributing to the correlations between vascular features and macular thickness. INL thickness was positively correlated with Width (r=0.122) and negatively correlated with LDR (r=−0.106). This might indicate that thinner INL was associated with a narrower diameter and elongated vessels relative to their diameter. Such a geometric pattern could potentially reflect vascular remodeling in response to altered metabolic demands or could be an early sign of microvascular dysfunction^32^. Previous studies have shown that changes in vessel morphology, including alterations in LDR, can be indicative of various retinal pathologies and systemic conditions such as diabetes and hypertension^33^. In addition, we found INL was associated with Vessel Area Density. This is consistent with the findings in a previous study^34^, which proposed the retinal vascular network had a more important role for the inner retina, while the oxygen and nutrition supply of the outer retina could rely on the choroid^27^.

The CCA revealed that the first three canonical dimensions explained more than 90% of the variance. The parameters that had strong correlations in the pairwise correlation analysis also primarily contributed to the correlations between the two sets of parameters. The first canonical dimension, explaining 43.8% of the variance, showed a moderate positive correlation (ρ = 0.265) between retinal vascular measurements and retinal layer thickness. The moderate strength of the correlation suggests that these two sets of measurements provide complementary rather than redundant information about retinal health and structure. This is partially evidenced by a previous study that reported that the combined use of CFP and OCT biomarkers led to improved performance for predicting late age-related macular degeneration development^35^. This highlights the value of the retinal vascular and neural layer as a composite biomarker. Further research should extend the application of grouped retinal vascular and neural biomarkers to systemic disease prediction. The information obtained on retinal vascular architecture and neural status especially may be relevant to changes in brain functions^36^ and potentially could serve as early biomarkers.

The bidirectional relationships revealed by Mendelian randomization analyses point to shared genetic influences between retinal vascular features and retinal layer thicknesses. This genetic overlap suggests common developmental pathways and regulatory mechanisms governing both vascular and neuronal components of the retina. A previous study reported that genes and signaling pathways, such as the Norrin signaling pathway mediated by the FZD4/LRP5/TSPAN12 receptor complex, involved in angiogenesis and neurogenesis during retinal development may have pleiotropic effects, influencing vascular architecture and neuronal layer formation^37^. Furthermore, the consistent associations across multiple retinal layers and vascular features indicate that these shared genetic factors may have broad effects on retinal structure and function. Understanding these shared genetic influences could provide valuable insights into the pathogenesis of retinal diseases and potentially facilitate the identification of new therapeutic targets.

Our study presents several strengths including large sample size, inclusion of a wide range of retinal vessel and neural layer features, and the employment of advanced statistical methods including CCA and Mendelian randomization. The bidirectional relationships suggest the presence of shared genetic influences and developmental pathways between vascular and neuronal components of the retina. This knowledge could lead to new therapeutic targets for conditions affecting both neurological and vascular health.

Despite its strengths, our study has some limitations. First, the analyses were performed in the population with a majority being Caucasians, which may affect the generalizability of the findings. Additionally, due to the low availability of GWAS data on OCT-derived retinal layers, we used data from a replication study in LIFE-Adult-study. Therefore, the SNPs included may not be all SNPs that can be identified in this study population. Additionally, the exclusion of eyes with IOP ≥21 mmHg or ≤8 mmHg may omit eyes at risk for or in the early stages of glaucoma, particularly those with normal-tension glaucoma. Finally, while the study provides a comprehensive view of retinal structure-vasculature relationships, it may not capture all the functional implications of these relationships in terms of visual performance or disease progression. The clinical implications of these results should be explored in future studies.

In conclusion, we revealed that macular thickness was associated with vascular Density and Caliber measurements. These results are mainly attributable to their associations with GC-IPL and INL thickness. Additionally, the multidimensional relationships revealed by CCA demonstrate the complementary nature of the two sets of parameters and highlight their value as a composite biomarker for both ocular and systemic conditions. Moreover, Mendelian Randomization uncovered a bidirectional relationship between retinal layer thicknesses and vascular features. This relationship offers insights that may be useful for developing novel therapeutic approaches targeting both vascular and neuronal components of the retina.

## Supporting information

STROBE_checklist_cross-sectional

## Data Availability

Data used in this study were from UK Biobank.

https://www.ukbiobank.ac.uk

