## Supplementary tables 1-7, Supplementary Text 1, and Supplementary figures 1-3 for "Relationship between Retinal Vessels and OCT-Derived Retinal Neural Parameters"

Supplementary Table 1. Definitions of variables

| **Variable** | **Definition and collection** | **Link** |
| --- | --- | --- |
| Age | Age when attended assessment centre | https://biobank.ndph.ox.ac.uk/showcase/field.cgi?id=21003 |
| Sex | Sex of participant. Acquired from central registry at recruitment, but in some cases updated by the participant. Hence this field may contain a mixture of the sex the NHS had recorded for the participant and self-reported sex. | http://biobank.ndph.ox.ac.uk/ukb/field.cgi?id=31 |
| Body mass index (BMI) | Defined as weight in kilogrammes divided by height in metres squared. BMI value here is constructed from height and weight measured during the initial Assessment Centre visit. Value is not present if either of these readings were omitted. | http://biobank.ndph.ox.ac.uk/ukb/field.cgi?id=21001 |
| Physical activity | Physical activity levels were determined by International Physical Activity Questionnaire (IPAQ). Three levels: Low/Moderate/High | http://biobank.ndph.ox.ac.uk/ukb/field.cgi?id=22032 |
| Diabetes | ACE touchscreen question \Has a doctor ever told you that you have diabetes?\" If the participant activated the Help button they were shown the message: If you are unsure if you have been told you had diabetes select Do not know and you will be asked about this by an interviewer later during this visit. " | http://biobank.ndph.ox.ac.uk/ukb/field.cgi?id=2443 |
| Smoking status | This field summarises the current/past smoking status of the participant. Only those who answered "Never" are considered non-smokers | http://biobank.ndph.ox.ac.uk/ukb/field.cgi?id=20116 |
| Intra-ocular pressure | Goldmann-correlated intraocular pressure. Units of measurement are mmHg. | https://biobank.ndph.ox.ac.uk/showcase/field.cgi?id=5263; https://biobank.ndph.ox.ac.uk/showcase/field.cgi?id=5255 |
| Low-Density Lipoprotein | Measured by enzyme immunoinhibition analysis on a Beckman Coulter AU5800. Units of measurement are mmol/L. | https://biobank.ndph.ox.ac.uk/showcase/field.cgi?id=30780 |
| High-Density Lipoprotein | Measured by enzyme immunoinhibition analysis on a Beckman Coulter AU5800. Units of measurement are mmol/L. | https://biobank.ndph.ox.ac.uk/showcase/field.cgi?id=30760 |
| Glycosylated hemoglobin | Measured by HPLC analysis on a Bio-Rad VARIANT II Turbo | https://biobank.ndph.ox.ac.uk/showcase/field.cgi?id=30750 |
| Systolic Blood Pressure | Blood pressure, automated reading, systolic. Omron device. Units of measurement are mmHg. Measured twice and the mean value was used. | https://biobank.ndph.ox.ac.uk/showcase/field.cgi?id=4080 |
| Diastolic Blood Pressure | Blood pressure, automated reading, diastolic. Omron device. Units of measurement are mmHg. Measured twice and the mean value was used. | https://biobank.ndph.ox.ac.uk/showcase/field.cgi?id=4079 |
| Cardiovascular Diseases | In addition, those reported vascular/heart problems at interview: ACE touchscreen question \Has a doctor ever told you that you have had any of the following conditions? | http://biobank.ndph.ox.ac.uk/ukb/field.cgi?id=6150; |
| Diabetic eye disease | Prior record of event was determined using data coding "1276" from Field ID 20002 for diabetic eye disease. In addition, those reported eye problems at interview: ACE touchscreen question \Has a doctor ever told you that you have had any of the following conditions? | https://biobank.ndph.ox.ac.uk/showcase/field.cgi?id=20002;  http://biobank.ndph.ox.ac.uk/ukb/field.cgi?id=6148; https://biobank.ndph.ox.ac.uk/showcase/coding.cgi?id=6 |
| Retinal Disease | Prior record of event was determined using data coding "1275, 1281, 1282 " from Field ID 20002 for retinal problem, retinal detachment, retinal artery/vein occlusion. In addition, those reported eye problems at interview: ACE touchscreen question \Has a doctor ever told you that you have had any of the following conditions? | https://biobank.ndph.ox.ac.uk/showcase/field.cgi?id=20002;  http://biobank.ndph.ox.ac.uk/ukb/field.cgi?id=6148; https://biobank.ndph.ox.ac.uk/showcase/coding.cgi?id=6 |
| Macular degeneration | Those reported eye problems at interview: ACE touchscreen question \Has a doctor ever told you that you have had any of the following conditions? | http://biobank.ndph.ox.ac.uk/ukb/field.cgi?id=6148; |
| Glaucoma | Prior record of event was determined using data coding "1277" from Field ID 20002 for glaucoma.  If participants received glaucoma surgery, decided using data code from Field 5326 and 5327, the eye was defined as glaucoma-affected.  In addition, those reported eye problems at interview: ACE touchscreen question \Has a doctor ever told you that you have had any of the following conditions?  The eye affected by glaucoma was decided using data code from Field 6119. | https://biobank.ndph.ox.ac.uk/showcase/field.cgi?id=20002;  http://biobank.ndph.ox.ac.uk/ukb/field.cgi?id=6148;  http://biobank.ndph.ox.ac.uk/ukb/field.cgi?id=6119; https://biobank.ndph.ox.ac.uk/showcase/coding.cgi?id=6 |

Supplementary Table 2. Definitions of measure types

| Category | Measure Type | Definition |
| --- | --- | --- |
| Tortuosity | Inflection Count Tortuosity | Calculated by counting the number of inflection points along the vessel. This method detects the inflection points of a given curve y=f(x) by applying a convolution to the y values and checking for changes in the sign of this convolution, each sign change is interpreted as an inflection point. |
|  | Fractal tortuosity | the Minkowski–Bouligand dimension of the vessel segment |
|  | Tortuosity density | this measure evaluates vessel tortuosity by summing local contributions to tortuosity by assessing how much each turn curve is different from a smooth curve |
|  | Curve angle | the mean segment angles between each branch sampled at a length of 10 pixels |
| Complexity | Number | number of vascular segments, or vascular trees, or bifurcation and branching points |
|  | Level | number of segments passing by an endpoint/branching/bifurcation point |
|  | Strahler | a numerical measure of its branching complexity using Strahler number |
| Density | Vessel Area Density | ratio of the area occupied by vessels divided by the total area of the 6*6 mm macular region |
|  | Vessel Skeleton Density | the ratio of the total length of vessels to the total area of the image at 1 pixel, a measure of overall vessel length within the 6*6 mm macular region |
|  | Branching Density | number of identified branchpoints divided by total vessel length |
|  | Bifurcation Density | number of identified bifurcations divided by total vessel length |
| Caliber | Mean width of all segments | mean value of diameters of all identified vessel segments within the 6*6 mm macular region |
|  | Length Diameter Ratio | the ratio of the length between two branching points to the trunk vessel width |
| Branching Angle | Asymmetry Ratio | (min (d1, d2)/max(d1/d2)) ^2, d1 and d2 denote diameters of the daughters of the root segment of vessel |
|  | Angular Asymmetry | the absolute difference between the angles of each daughter vessel with the root vessel |
|  | Branching Coefficient | (d1+d2) ^2/d0^2, d0 = parent vessel diameter (px), d1 = daughter's vessel diameter (px), d2 = diameter of daughter vessel two (μm), it explains the relationship between the calibres of the parent vessel compared to the daughter vessels at a bifurcation. |
|  | Junctional Exponent Deviation | the extent to which the relationship between the diameter of the parent vessel and the daughter vessels deviates from theoretically defined optimum (Murray's law). |
|  | Branching Angle | the angles between the sampled centreline of the root segments of vessel and its daughter segments s1 and s2 near the branching point. Murray proposed in 1926, the optimal arteriolar branching angle to be 75 degrees, and any deviation from the optimal angle was seen as less optimal for the retinal circulatory system. |

Supplementary Text 1. Data Sources for Mendelian Randomization

A) GWAS data of retinal Vessel Density and Fractal Dimension obtained in the UK Biobank Study

SNPs related to retinal vessel density and fractal dimension were obtained from the UK Biobank. The vascular network in 97,895 photographs from 54,813 participants was segmented with U-Nets on the Google Cloud. Two main vascular indicators were assessed: vascular density and branching complexity, which were determined by fractal dimension (FD). Vascular branching complexity was measured by calculating the FD using a box-counting method. On the other hand, Vascular density was defined as the total number of segmented pixels, normalized to a 320x320 pixel dimension per image. To find the genotypes impacting these vascular indices, they subsequently performed genome-wide association analyses of retinal FD and vascular density. Hail-0.2 software was used to conduct GWASs on 38 932 unrelated people and 15 580 782 variations with a minor allele frequency >0.001 on Google Cloud.

B) GWAS data of retinal layer thickness obtained in the LIFE-Adult Study

SNPs associated with OCT layer thickness were obtained from a replication study using the LIFE-Adult Study, which comprised 6,313 individuals of central European descent from Germany. Topcon Advanced Boundary Segmentation was used for automatic segmentation of the inner and outer retinal boundaries and retinal sublayers. Genotyping was performed with Affymetrix Axiom CEU1 and genotyping calling software Affymetrix Power Tools v1.20.6. The OCT retinal layers used in the GWAS study included ganglion cell complex (GCC), retinal nerve fibre layer (RNFL), ganglion cell layer (GCL), inner plexiform layer (IPL), inner nuclear layer (INL), outer plexiform layer (OPL), photoreceptor segments (PS) layer, retinal pigment epithelium–Bruch's membrane complex (RPE-BM), and choroid-scleral interface (CSI). GCC included RNFL+GCL + IPL. PS layer included external limiting membrane- inner segment/outer segment (ISOS) + ISOS-retinal pigment epithelium.

Supplementary Table 3. Correlations between retinal vascular features and retinal layer thickness

|  | Macular thickness  Coefficient (p) | RNFL  Coefficient (p) | GCIPL  Coefficient (p) | INL-RPE  Coefficient (p) | INL  Coefficient (p) | ELM-INL  Coefficient (p) | ISOS-ELM  Coefficient (p) | RPE-ISOS  Coefficient (p) | RPE  Coefficient (p) |
| --- | --- | --- | --- | --- | --- | --- | --- | --- | --- |
| **Tortuosity Density** | 0.038 [<0.001] | 0.018 [<0.001] | 0.053 [<0.001] | 0.001 [0.811] | 0.043 [<0.001] | 0.015 [<0.001] | -0.032 [<0.001] | -0.013 [<0.001] | 0.009 [0.018] |
| **Inflection Count Tortuosity** | 0.050 [<0.001] | 0.015 [<0.001] | 0.075 [<0.001] | 0.004 [0.270] | 0.057 [<0.001] | 0.017 [<0.001] | -0.027 [<0.001] | -0.010 [0.012] | 0.017 [<0.001] |
| **Fractal Tortuosity** | -0.029 [<0.001] | -0.004 [0.345] | -0.041 [<0.001] | -0.008 [0.050] | -0.028 [<0.001] | -0.007 [0.061] | -0.005 [0.159] | 0.002 [0.663] | -0.021 [<0.001] |
| **Curve Angle** | 0.057 [<0.001] | 0.057 [<0.001] | 0.041 [<0.001] | 0.025 [<0.001] | 0.014 [<0.001] | 0.025 [<0.001] | -0.000 [0.916] | 0.008 [0.034] | 0.002 [0.562] |
| **Vessel Skeleton Density (vein)** | 0.102 [<0.001] | 0.064 [<0.001] | 0.135 [<0.001] | 0.014 [<0.001] | 0.072 [<0.001] | 0.001 [0.820] | 0.038 [<0.001] | 0.009 [0.018] | 0.038 [<0.001] |
| **Vessel Skeleton Density (artery)** | 0.132 [<0.001] | 0.065 [<0.001] | 0.170 [<0.001] | 0.026 [<0.001] | 0.101 [<0.001] | 0.008 [0.045] | 0.055 [<0.001] | 0.022 [<0.001] | 0.034 [<0.001] |
| **Vessel Area Density (vein)** | 0.116 [<0.001] | 0.046 [<0.001] | 0.152 [<0.001] | 0.029 [<0.001] | 0.095 [<0.001] | 0.021 [<0.001] | 0.023 [<0.001] | 0.011 [0.003] | 0.029 [<0.001] |
| **Vessel Area Density (artery)** | 0.161 [<0.001] | 0.059 [<0.001] | 0.199 [<0.001] | 0.052 [<0.001] | 0.127 [<0.001] | 0.036 [<0.001] | 0.053 [<0.001] | 0.025 [<0.001] | 0.030 [<0.001] |
| **Branching Density** | 0.064 [<0.001] | -0.003 [0.381] | 0.082 [<0.001] | 0.036 [<0.001] | 0.049 [<0.001] | 0.038 [<0.001] | 0.014 [<0.001] | 0.001 [0.720] | 0.034 [<0.001] |
| **Bifurcation Density** | 0.061 [<0.001] | 0.020 [<0.001] | 0.071 [<0.001] | 0.023 [<0.001] | 0.051 [<0.001] | 0.025 [<0.001] | 0.008 [0.030] | 0.001 [0.810] | 0.025 [<0.001] |
| **Strahler** | 0.051 [<0.001] | 0.009 [0.020] | 0.086 [<0.001] | 0.002 [0.585] | 0.052 [<0.001] | 0.003 [0.509] | -0.001 [0.871] | 0.001 [0.705] | 0.016 [<0.001] |
| **Number of Trees** | 0.005 [0.227] | -0.002 [0.599] | 0.009 [0.020] | 0.001 [0.840] | -0.006 [0.120] | 0.008 [0.050] | 0.019 [<0.001] | -0.009 [0.014] | 0.002 [0.516] |
| **Number of Segments** | 0.078 [<0.001] | -0.032 [<0.001] | 0.175 [<0.001] | 0.001 [0.795] | 0.077 [<0.001] | -0.006 [0.099] | 0.014 [<0.001] | 0.001 [0.760] | 0.055 [<0.001] |
| **Number of Branching** | 0.073 [<0.001] | -0.029 [<0.001] | 0.157 [<0.001] | 0.006 [0.134] | 0.070 [<0.001] | -0.003 [0.435] | 0.014 [<0.001] | 0.005 [0.190] | 0.047 [<0.001] |
| **Number of Bifurcation** | 0.050 [<0.001] | -0.029 [<0.001] | 0.124 [<0.001] | -0.002 [0.552] | 0.051 [<0.001] | -0.011 [0.006] | 0.010 [0.012] | 0.002 [0.652] | 0.039 [<0.001] |
| **Level** | 0.056 [<0.001] | 0.013 [<0.001] | 0.085 [<0.001] | 0.010 [0.009] | 0.053 [<0.001] | 0.009 [0.020] | 0.010 [0.009] | 0.004 [0.349] | 0.016 [<0.001] |
| **Width** | 0.118 [<0.001] | -0.023 [<0.001] | 0.131 [<0.001] | 0.084 [<0.001] | 0.122 [<0.001] | 0.093 [<0.001] | -0.012 [0.002] | 0.015 [<0.001] | -0.004 [0.334] |
| **Length Diameter Ratio** | -0.119 [<0.001] | 0.001 [0.807] | -0.143 [<0.001] | -0.067 [<0.001] | -0.106 [<0.001] | -0.072 [<0.001] | -0.004 [0.249] | -0.009 [0.019] | -0.034 [<0.001] |
| **Junctional Exponent Deviation** | 0.046 [<0.001] | -0.021 [<0.001] | 0.058 [<0.001] | 0.034 [<0.001] | 0.050 [<0.001] | 0.043 [<0.001] | 0.004 [0.242] | -0.006 [0.099] | 0.010 [0.009] |
| **Branching Coefficient** | -0.040 [<0.001] | 0.010 [0.011] | -0.040 [<0.001] | -0.034 [<0.001] | -0.037 [<0.001] | -0.034 [<0.001] | -0.010 [0.010] | -0.008 [0.033] | 0.001 [0.727] |
| **Branching Angle Measure** | 0.028 [<0.001] | 0.034 [<0.001] | 0.017 [<0.001] | 0.009 [0.019] | 0.008 [0.031] | 0.013 [<0.001] | -0.021 [<0.001] | 0.005 [0.214] | -0.010 [0.013] |
| **Asymmetry Ratio** | 0.047 [<0.001] | 0.026 [<0.001] | 0.041 [<0.001] | 0.026 [<0.001] | 0.036 [<0.001] | 0.029 [<0.001] | -0.007 [0.068] | 0.002 [0.526] | 0.008 [0.033] |
| **Angular Asymmetry** | 0.031 [<0.001] | 0.048 [<0.001] | 0.039 [<0.001] | -0.017 [<0.001] | 0.026 [<0.001] | -0.009 [0.015] | 0.005 [0.221] | -0.015 [<0.001] | 0.013 [<0.001] |

Notes: RNFL = Retinal Nerve Fiber Layer, GCIPL = Ganglion Cell-Inner Plexiform Layer, INL = Inner Nuclear Layer, ELM = External Limiting Membrane, ISOS = Inner Segment/Outer Segment junction, RPE = Retinal Pigment Epithelium Correlation coefficients are rounded to 3 decimal places. P-values are shown in square brackets, rounded to 3 decimal places or shown as <0.001 if smaller than 0.001.

Supplementary Table 4. Correlations between retinal vascular features and retinal layer thickness by sex

Table 4A. Correlations between retinal vascular features and retinal layer thickness in males

|  | Macular thickness  Coefficient (p) | RNFL  Coefficient (p) | GCIPL  Coefficient (p) | INL-RPE  Coefficient (p) | INL  Coefficient (p) | ELM-INL  Coefficient (p) | ISOS-ELM  Coefficient (p) | RPE-ISOS  Coefficient (p) | RPE  Coefficient (p) |
| --- | --- | --- | --- | --- | --- | --- | --- | --- | --- |
| **Tortuosity Density** | 0.036 [<0.001] | 0.015 [0.009] | 0.050 [<0.001] | 0.001 [0.903] | 0.039 [<0.001] | 0.014 [0.012] | -0.028 [<0.001] | -0.015 [0.007] | 0.017 [0.002] |
| **Inflection Count Tortuosity** | 0.049 [<0.001] | 0.017 [0.004] | 0.074 [<0.001] | 0.015 [0.009] | 0.048 [<0.001] | 0.015 [0.009] | -0.029 [<0.001] | -0.007 [0.237] | 0.020 [0.001] |
| **Fractal Tortuosity** | -0.039 [<0.001] | -0.009 [0.136] | -0.049 [<0.001] | -0.016 [0.007] | -0.032 [<0.001] | -0.016 [0.007] | 0.000 [0.983] | -0.005 [0.401] | -0.014 [0.015] |
| **Curve Angle** | 0.062 [<0.001] | 0.058 [<0.001] | 0.045 [<0.001] | 0.028 [<0.001] | 0.019 [0.001] | 0.027 [<0.001] | 0.007 [0.210] | 0.009 [0.133] | 0.008 [0.154] |
| **Vessel Skeleton Density (vein)** | 0.117 [<0.001] | 0.065 [<0.001] | 0.157 [<0.001] | 0.022 [0.000] | 0.082 [<0.001] | 0.014 [0.012] | 0.024 [<0.001] | 0.008 [0.191] | 0.030 [<0.001] |
| **Vessel Skeleton Density (artery)** | 0.149 [<0.001] | 0.063 [<0.001] | 0.193 [<0.001] | 0.036 [<0.001] | 0.113 [<0.001] | 0.024 [<0.001] | 0.039 [<0.001] | 0.022 [0.000] | 0.037 [<0.001] |
| **Vessel Area Density (vein)** | 0.122 [<0.001] | 0.045 [<0.001] | 0.166 [<0.001] | 0.029 [<0.001] | 0.099 [<0.001] | 0.028 [<0.001] | 0.004 [0.487] | 0.008 [0.148] | 0.025 [<0.001] |
| **Vessel Area Density (artery)** | 0.176 [<0.001] | 0.059 [<0.001] | 0.217 [<0.001] | 0.061 [<0.001] | 0.138 [<0.001] | 0.053 [<0.001] | 0.036 [<0.001] | 0.023 [<0.001] | 0.036 [<0.001] |
| **Branching Density** | 0.075 [<0.001] | 0.006 [0.316] | 0.088 [<0.001] | 0.042 [<0.001] | 0.052 [<0.001] | 0.044 [<0.001] | 0.007 [0.248] | 0.010 [0.098] | 0.023 [<0.001] |
| **Bifurcation Density** | 0.068 [<0.001] | 0.021 [0.000] | 0.082 [<0.001] | 0.026 [<0.001] | 0.057 [<0.001] | 0.026 [<0.001] | -0.005 [0.399] | 0.005 [0.408] | 0.022 [0.000] |
| **Strahler** | 0.048 [<0.001] | 0.002 [0.779] | 0.088 [<0.001] | 0.000 [0.969] | 0.051 [<0.001] | 0.000 [0.969] | -0.005 [0.415] | 0.003 [0.575] | 0.018 [0.002] |
| **Number of Trees** | 0.002 [0.747] | -0.005 [0.396] | 0.008 [0.165] | -0.003 [0.555] | -0.005 [0.342] | 0.002 [0.669] | 0.029 [<0.001] | -0.010 [0.092] | 0.006 [0.262] |
| **Number of Segments** | 0.089 [<0.001] | -0.034 [<0.001] | 0.189 [<0.001] | 0.004 [0.484] | 0.081 [<0.001] | 0.001 [0.856] | -0.004 [0.500] | 0.007 [0.212] | 0.050 [<0.001] |
| **Number of Branching** | 0.082 [<0.001] | -0.031 [<0.001] | 0.170 [<0.001] | 0.006 [0.322] | 0.072 [<0.001] | 0.002 [0.787] | -0.005 [0.375] | 0.008 [0.178] | 0.043 [<0.001] |
| **Number of Bifurcation** | 0.064 [<0.001] | -0.031 [<0.001] | 0.138 [<0.001] | 0.003 [0.603] | 0.059 [<0.001] | 0.003 [0.603] | 0.001 [0.927] | 0.008 [0.177] | 0.039 [<0.001] |
| **Level** | 0.053 [<0.001] | 0.011 [0.050] | 0.085 [<0.001] | 0.005 [0.427] | 0.049 [<0.001] | 0.005 [0.427] | 0.001 [0.855] | 0.003 [0.592] | 0.016 [0.005] |
| **Width** | 0.104 [<0.001] | -0.024 [<0.001] | 0.116 [<0.001] | 0.071 [<0.001] | 0.115 [<0.001] | 0.086 [<0.001] | -0.025 [<0.001] | 0.008 [0.143] | 0.002 [0.740] |
| **Length Diameter Ratio** | -0.127 [<0.001] | -0.005 [0.369] | -0.147 [<0.001] | -0.069 [<0.001] | -0.108 [<0.001] | -0.076 [<0.001] | 0.011 [0.061] | -0.014 [0.018] | -0.025 [<0.001] |
| **Junctional Exponent Deviation** | 0.049 [<0.001] | -0.023 [<0.001] | 0.064 [<0.001] | 0.033 [<0.001] | 0.054 [<0.001] | 0.048 [<0.001] | -0.006 [0.300] | -0.014 [0.016] | 0.018 [0.002] |
| **Branching Coefficient** | -0.043 [<0.001] | 0.011 [0.057] | -0.045 [<0.001] | -0.036 [<0.001] | -0.038 [<0.001] | -0.036 [<0.001] | -0.003 [0.618] | -0.005 [0.352] | -0.002 [0.752] |
| **Branching Angle Measure** | 0.020 [0.001] | 0.028 [<0.001] | 0.015 [0.010] | 0.001 [0.876] | 0.006 [0.324] | 0.003 [0.641] | -0.029 [<0.001] | 0.008 [0.145] | -0.010 [0.078] |
| **Asymmetry Ratio** | 0.036 [<0.001] | 0.015 [0.008] | 0.037 [<0.001] | 0.024 [<0.001] | 0.030 [<0.001] | 0.024 [<0.001] | -0.017 [0.004] | -0.006 [0.265] | 0.009 [0.106] |
| **Angular Asymmetry** | 0.031 [<0.001] | 0.042 [<0.001] | 0.046 [<0.001] | -0.009 [0.103] | 0.026 [<0.001] | -0.004 [0.521] | -0.004 [0.521] | -0.011 [0.055] | 0.016 [0.004] |

Table 4B. Correlations between retinal vascular features and retinal layer thickness in females

|  | Macular thickness  Coefficient (p) | RNFL  Coefficient (p) | GCIPL  Coefficient (p) | INL-RPE  Coefficient (p) | INL  Coefficient (p) | ELM-INL  Coefficient (p) | ISOS-ELM  Coefficient (p) | RPE-ISOS  Coefficient (p) | RPE  Coefficient (p) |
| --- | --- | --- | --- | --- | --- | --- | --- | --- | --- |
| **Tortuosity Density** | 0.043 (<0.001) | 0.016 (0.002) | 0.055 (<0.001) | 0.008 (0.130) | 0.052 (<0.001) | 0.022 (<0.001) | -0.033 (<0.001) | -0.010 (0.059) | 0.008 (0.128) |
| **Inflection Count Tortuosity** | 0.051 (<0.001) | 0.014 (0.008) | 0.076 (<0.001) | 0.004 (0.482) | 0.064 (<0.001) | 0.018 (<0.001) | -0.026 (<0.001) | -0.012 (0.023) | 0.016 (0.002) |
| **Fractal Tortuosity** | -0.017 (0.001) | -0.004 (0.410) | -0.035 (<0.001) | 0.006 (0.206) | -0.018 (0.001) | 0.007 (0.159) | -0.007 (0.170) | 0.009 (0.087) | -0.019 (<0.001) |
| **Curve Angle** | 0.058 (<0.001) | 0.051 (<0.001) | 0.037 (<0.001) | 0.032 (<0.001) | 0.019 (<0.001) | 0.033 (<0.001) | -0.003 (0.500) | 0.010 (0.054) | 0.004 (0.422) |
| **Vessel Skeleton Density (vein)** | 0.084 (<0.001) | 0.072 (<0.001) | 0.117 (<0.001) | -0.006 (0.279) | 0.052 (<0.001) | -0.023 (<0.001) | 0.046 (<0.001) | 0.007 (0.172) | 0.034 (<0.001) |
| **Vessel Skeleton Density (artery)** | 0.118 (<0.001) | 0.067 (<0.001) | 0.152 (<0.001) | 0.019 (<0.001) | 0.092 (<0.001) | -0.005 (0.376) | 0.069 (<0.001) | 0.022 (<0.001) | 0.032 (<0.001) |
| **Vessel Area Density (vein)** | 0.106 (<0.001) | 0.054 (<0.001) | 0.141 (<0.001) | 0.018 (<0.001) | 0.081 (<0.001) | 0.005 (0.318) | 0.034 (<0.001) | 0.011 (0.038) | 0.024 (<0.001) |
| **Vessel Area Density (artery)** | 0.152 (<0.001) | 0.058 (<0.001) | 0.184 (<0.001) | 0.051 (<0.001) | 0.124 (<0.001) | 0.029 (<0.001) | 0.070 (<0.001) | 0.029 (<0.001) | 0.029 (<0.001) |
| **Branching Density** | 0.053 (<0.001) | -0.008 (0.129) | 0.077 (<0.001) | 0.027 (<0.001) | 0.043 (<0.001) | 0.030 (<0.001) | 0.017 (0.001) | -0.006 (0.208) | 0.038 (<0.001) |
| **Bifurcation Density** | 0.054 (<0.001) | 0.022 (<0.001) | 0.061 (<0.001) | 0.018 (<0.001) | 0.043 (<0.001) | 0.020 (<0.001) | 0.017 (0.001) | -0.003 (0.557) | 0.024 (<0.001) |
| **Strahler** | 0.055 (<0.001) | 0.013 (0.009) | 0.085 (<0.001) | 0.007 (0.159) | 0.056 (<0.001) | 0.008 (0.141) | 0.004 (0.475) | 0.001 (0.893) | 0.016 (0.001) |
| **Number of Trees** | 0.009 (0.094) | -0.002 (0.756) | 0.010 (0.057) | 0.008 (0.141) | -0.003 (0.530) | 0.015 (0.004) | 0.012 (0.015) | -0.008 (0.112) | 0.002 (0.708) |
| **Number of Segments** | 0.067 (<0.001) | -0.027 (<0.001) | 0.163 (<0.001) | -0.006 (0.242) | 0.069 (<0.001) | -0.017 (0.001) | 0.027 (<0.001) | -0.005 (0.355) | 0.055 (<0.001) |
| **Number of Branching** | 0.063 (<0.001) | -0.024 (<0.001) | 0.147 (<0.001) | -0.001 (0.842) | 0.063 (<0.001) | -0.013 (0.011) | 0.027 (<0.001) | 0.001 (0.802) | 0.045 (<0.001) |
| **Number of Bifurcation** | 0.039 (<0.001) | -0.026 (<0.001) | 0.113 (<0.001) | -0.011 (0.030) | 0.043 (<0.001) | -0.023 (<0.001) | 0.016 (0.001) | -0.003 (0.512) | 0.038 (<0.001) |
| **Level** | 0.058 (<0.001) | 0.015 (0.003) | 0.084 (<0.001) | 0.014 (0.007) | 0.056 (<0.001) | 0.011 (0.025) | 0.017 (0.001) | 0.004 (0.483) | 0.015 (0.003) |
| **Width** | 0.131 (<0.001) | -0.024 (<0.001) | 0.142 (<0.001) | 0.101 (<0.001) | 0.134 (<0.001) | 0.105 (<0.001) | 0.000 (0.944) | 0.022 (<0.001) | -0.005 (0.357) |
| **Length Diameter Ratio** | -0.112 (<0.001) | 0.003 (0.501) | -0.140 (<0.001) | -0.063 (<0.001) | -0.103 (<0.001) | -0.066 (<0.001) | -0.015 (0.003) | -0.004 (0.385) | -0.038 (<0.001) |
| **Junctional Exponent Deviation** | 0.044 (<0.001) | -0.019 (<0.001) | 0.053 (<0.001) | 0.034 (<0.001) | 0.046 (<0.001) | 0.038 (<0.001) | 0.012 (0.015) | -0.000 (0.946) | 0.004 (0.484) |
| **Branching Coefficient** | -0.035 (<0.001) | 0.007 (0.186) | -0.035 (<0.001) | -0.030 (<0.001) | -0.034 (<0.001) | -0.028 (<0.001) | -0.014 (0.007) | -0.009 (0.064) | 0.007 (0.196) |
| **Branching Angle Measure** | 0.038 (<0.001) | 0.036 (<0.001) | 0.018 (0.001) | 0.021 (<0.001) | 0.016 (0.002) | 0.026 (<0.001) | -0.013 (0.012) | 0.003 (0.550) | -0.005 (0.298) |
| **Asymmetry Ratio** | 0.054 (<0.001) | 0.037 (<0.001) | 0.043 (<0.001) | 0.028 (<0.001) | 0.038 (<0.001) | 0.027 (<0.001) | -0.001 (0.810) | 0.008 (0.109) | 0.003 (0.566) |
| **Angular Asymmetry** | 0.033 (<0.001) | 0.052 (<0.001) | 0.033 (<0.001) | -0.015 (0.003) | 0.029 (<0.001) | -0.007 (0.198) | 0.012 (0.017) | -0.018 (0.001) | 0.012 (0.018) |

Supplementary Table 5. Correlations between retinal vascular features and retinal layer thickness by age groups

Table 5A. Correlations between retinal vascular features and retinal layer thickness in participants aged 50 and under

|  | Macular thickness  Coefficient (p) | RNFL  Coefficient (p) | GCIPL  Coefficient (p) | INL-RPE  Coefficient (p) | INL  Coefficient (p) | ELM-INL  Coefficient (p) | ISOS-ELM  Coefficient (p) | RPE-ISOS  Coefficient (p) | RPE  Coefficient (p) |
| --- | --- | --- | --- | --- | --- | --- | --- | --- | --- |
| **Tortuosity Density** | 0.048 (<0.001) | 0.042 (<0.001) | 0.066 (<0.001) | -0.010 (0.122) | 0.050 (<0.001) | 0.012 (0.068) | -0.030 (<0.001) | -0.025 (<0.001) | 0.021 (0.001) |
| **Inflection Count Tortuosity** | 0.049 (<0.001) | 0.033 (<0.001) | 0.078 (<0.001) | -0.009 (0.159) | 0.056 (<0.001) | 0.014 (0.043) | -0.039 (<0.001) | -0.022 (0.001) | 0.022 (0.001) |
| **Fractal Tortuosity** | -0.001 (0.898) | 0.009 (0.193) | -0.011 (0.102) | 0.002 (0.718) | -0.016 (0.020) | -0.008 (0.221) | 0.012 (0.073) | 0.014 (0.034) | -0.017 (0.010) |
| **Curve Angle** | 0.057 (<0.001) | 0.072 (<0.001) | 0.030 (<0.001) | 0.026 (<0.001) | 0.016 (0.014) | 0.033 (<0.001) | 0.004 (0.598) | 0.002 (0.724) | 0.004 (0.574) |
| **Vessel Skeleton Density (vein)** | 0.085 (<0.001) | 0.068 (<0.001) | 0.091 (<0.001) | 0.020 (0.002) | 0.071 (<0.001) | 0.024 (<0.001) | -0.011 (0.091) | 0.007 (0.326) | 0.004 (0.594) |
| **Vessel Skeleton Density (artery)** | 0.096 (<0.001) | 0.056 (<0.001) | 0.117 (<0.001) | 0.016 (0.015) | 0.087 (<0.001) | 0.020 (0.003) | -0.007 (0.281) | 0.011 (0.102) | -0.007 (0.299) |
| **Vessel Area Density (vein)** | 0.108 (<0.001) | 0.053 (<0.001) | 0.120 (<0.001) | 0.036 (<0.001) | 0.102 (<0.001) | 0.043 (<0.001) | -0.014 (0.033) | 0.009 (0.196) | -0.000 (0.993) |
| **Vessel Area Density (artery)** | 0.137 (<0.001) | 0.058 (<0.001) | 0.161 (<0.001) | 0.046 (<0.001) | 0.118 (<0.001) | 0.052 (<0.001) | 0.001 (0.872) | 0.014 (0.035) | -0.008 (0.216) |
| **Branching Density** | 0.027 (<0.001) | -0.016 (0.015) | 0.039 (<0.001) | 0.020 (0.003) | 0.034 (<0.001) | 0.041 (<0.001) | -0.018 (0.008) | -0.017 (0.013) | 0.020 (0.003) |
| **Bifurcation Density** | 0.037 (<0.001) | 0.013 (0.049) | 0.039 (<0.001) | 0.016 (0.021) | 0.036 (<0.001) | 0.031 (<0.001) | -0.008 (0.241) | -0.012 (0.078) | 0.022 (0.001) |
| **Strahler** | 0.042 (<0.001) | 0.006 (0.342) | 0.076 (<0.001) | -0.006 (0.387) | 0.045 (<0.001) | 0.003 (0.616) | -0.010 (0.144) | -0.008 (0.232) | 0.007 (0.296) |
| **Number of Trees** | 0.009 (0.186) | 0.001 (0.923) | 0.016 (0.017) | -0.002 (0.718) | 0.007 (0.276) | 0.005 (0.503) | 0.011 (0.098) | -0.011 (0.091) | 0.014 (0.042) |
| **Number of Segments** | 0.023 (0.001) | -0.065 (<0.001) | 0.120 (<0.001) | -0.023 (0.001) | 0.057 (<0.001) | 0.005 (0.438) | -0.052 (<0.001) | -0.032 (<0.001) | 0.033 (<0.001) |
| **Number of Branching** | 0.024 (<0.001) | -0.056 (<0.001) | 0.105 (<0.001) | -0.014 (0.036) | 0.050 (<0.001) | 0.005 (0.448) | -0.041 (<0.001) | -0.021 (0.001) | 0.023 (<0.001) |
| **Number of Bifurcation** | 0.009 (0.159) | -0.051 (<0.001) | 0.076 (<0.001) | -0.018 (0.007) | 0.039 (<0.001) | -0.001 (0.901) | -0.043 (<0.001) | -0.019 (0.004) | 0.021 (0.001) |
| **Level** | 0.052 (<0.001) | 0.018 (0.008) | 0.078 (<0.001) | 0.004 (0.518) | 0.046 (<0.001) | 0.009 (0.186) | -0.005 (0.434) | 0.001 (0.883) | 0.004 (0.529) |
| **Width** | 0.137 (<0.001) | 0.002 (0.814) | 0.149 (<0.001) | 0.086 (<0.001) | 0.126 (<0.001) | 0.093 (<0.001) | 0.003 (0.691) | 0.021 (0.001) | -0.012 (0.063) |
| **Length Diameter Ratio** | -0.086 (<0.001) | 0.009 (0.201) | -0.102 (<0.001) | -0.052 (<0.001) | -0.091 (<0.001) | -0.079 (<0.001) | 0.026 (<0.001) | 0.012 (0.082) | -0.023 (0.001) |
| **Junctional Exponent Deviation** | 0.049 (<0.001) | -0.027 (<0.001) | 0.063 (<0.001) | 0.045 (<0.001) | 0.054 (<0.001) | 0.054 (<0.001) | 0.007 (0.310) | 0.004 (0.526) | 0.005 (0.424) |
| **Branching Coefficient** | -0.047 (<0.001) | 0.015 (0.021) | -0.046 (<0.001) | -0.049 (<0.001) | -0.042 (<0.001) | -0.043 (<0.001) | -0.014 (0.043) | -0.025 (<0.001) | 0.013 (0.051) |
| **Branching Angle Measure** | 0.032 (<0.001) | 0.052 (<0.001) | 0.016 (0.015) | -0.002 (0.766) | 0.010 (0.118) | 0.010 (0.151) | -0.014 (0.038) | -0.011 (0.105) | 0.002 (0.781) |
| **Asymmetry Ratio** | 0.041 (<0.001) | 0.037 (<0.001) | 0.035 (<0.001) | 0.015 (0.026) | 0.026 (<0.001) | 0.021 (0.002) | -0.010 (0.152) | -0.000 (0.956) | 0.009 (0.191) |
| **Angular Asymmetry** | 0.031 (<0.001) | 0.065 (<0.001) | 0.033 (<0.001) | -0.022 (0.001) | 0.019 (0.005) | -0.018 (0.007) | 0.005 (0.460) | -0.015 (0.026) | 0.001 (0.910) |

Table 5B. Correlations between retinal vascular features and retinal layer thickness in participants aged 50 to 60

|  | Macular thickness  Coefficient (p) | RNFL  Coefficient (p) | GCIPL  Coefficient (p) | INL-RPE  Coefficient (p) | INL  Coefficient (p) | ELM-INL  Coefficient (p) | ISOS-ELM  Coefficient (p) | RPE-ISOS  Coefficient (p) | RPE  Coefficient (p) |
| --- | --- | --- | --- | --- | --- | --- | --- | --- | --- |
| **Tortuosity Density** | 0.033 (<0.001) | 0.008 (0.190) | 0.050 (<0.001) | 0.013 (0.046) | 0.044 (<0.001) | 0.013 (0.046) | -0.026 (<0.001) | -0.018 (0.006) | 0.005 (0.432) |
| **Inflection Count Tortuosity** | 0.048 (<0.001) | 0.007 (0.247) | 0.066 (<0.001) | 0.020 (0.002) | 0.060 (<0.001) | 0.020 (0.002) | -0.025 (<0.001) | -0.009 (0.180) | 0.011 (0.097) |
| **Fractal Tortuosity** | -0.039 (<0.001) | -0.000 (0.972) | -0.037 (<0.001) | -0.028 (<0.001) | -0.038 (<0.001) | -0.028 (<0.001) | 0.004 (0.503) | -0.010 (0.118) | -0.012 (0.067) |
| **Curve Angle** | 0.058 (<0.001) | 0.052 (<0.001) | 0.046 (<0.001) | 0.025 (<0.001) | 0.014 (0.028) | 0.025 (<0.001) | -0.003 (0.623) | 0.004 (0.556) | -0.003 (0.667) |
| **Vessel Skeleton Density (vein)** | 0.086 (<0.001) | 0.054 (<0.001) | 0.098 (<0.001) | 0.020 (0.002) | 0.064 (<0.001) | 0.012 (0.056) | -0.010 (0.120) | 0.016 (0.014) | 0.014 (0.031) |
| **Vessel Skeleton Density (artery)** | 0.129 (<0.001) | 0.064 (<0.001) | 0.139 (<0.001) | 0.045 (<0.001) | 0.103 (<0.001) | 0.030 (<0.001) | 0.005 (0.458) | 0.037 (<0.001) | -0.006 (0.328) |
| **Vessel Area Density (vein)** | 0.109 (<0.001) | 0.034 (<0.001) | 0.131 (<0.001) | 0.040 (<0.001) | 0.093 (<0.001) | 0.037 (<0.001) | -0.012 (0.061) | 0.018 (0.005) | 0.011 (0.077) |
| **Vessel Area Density (artery)** | 0.159 (<0.001) | 0.057 (<0.001) | 0.167 (<0.001) | 0.072 (<0.001) | 0.131 (<0.001) | 0.059 (<0.001) | 0.008 (0.225) | 0.042 (<0.001) | -0.005 (0.398) |
| **Branching Density** | 0.064 (<0.001) | -0.011 (0.083) | 0.067 (<0.001) | 0.049 (<0.001) | 0.050 (<0.001) | 0.054 (<0.001) | -0.016 (0.014) | 0.011 (0.095) | 0.013 (0.041) |
| **Bifurcation Density** | 0.058 (<0.001) | 0.024 (<0.001) | 0.059 (<0.001) | 0.026 (<0.001) | 0.051 (<0.001) | 0.033 (<0.001) | -0.020 (0.002) | 0.002 (0.764) | 0.006 (0.349) |
| **Strahler** | 0.053 (<0.001) | 0.011 (0.082) | 0.085 (<0.001) | 0.005 (0.448) | 0.058 (<0.001) | 0.010 (0.118) | -0.018 (0.004) | -0.000 (0.955) | 0.017 (0.007) |
| **Number of Trees** | 0.012 (0.073) | -0.003 (0.601) | 0.007 (0.247) | 0.016 (0.014) | -0.002 (0.761) | 0.022 (<0.001) | 0.017 (0.010) | -0.003 (0.658) | -0.009 (0.143) |
| **Number of Segments** | 0.070 (<0.001) | -0.041 (<0.001) | 0.142 (<0.001) | 0.017 (0.008) | 0.079 (<0.001) | 0.012 (0.059) | -0.038 (<0.001) | 0.016 (0.011) | 0.021 (0.001) |
| **Number of Branching** | 0.065 (<0.001) | -0.036 (<0.001) | 0.126 (<0.001) | 0.018 (0.006) | 0.070 (<0.001) | 0.012 (0.056) | -0.030 (<0.001) | 0.016 (0.012) | 0.021 (0.001) |
| **Number of Bifurcation** | 0.043 (<0.001) | -0.037 (<0.001) | 0.098 (<0.001) | 0.012 (0.061) | 0.051 (<0.001) | 0.005 (0.431) | -0.030 (<0.001) | 0.014 (0.029) | 0.010 (0.140) |
| **Level** | 0.056 (<0.001) | 0.012 (0.069) | 0.078 (<0.001) | 0.014 (0.031) | 0.059 (<0.001) | 0.016 (0.013) | 0.004 (0.499) | 0.000 (0.957) | 0.017 (0.009) |
| **Width** | 0.124 (<0.001) | -0.031 (<0.001) | 0.142 (<0.001) | 0.085 (<0.001) | 0.132 (<0.001) | 0.096 (<0.001) | -0.004 (0.556) | 0.013 (0.046) | 0.007 (0.256) |
| **Length Diameter Ratio** | -0.122 (<0.001) | 0.006 (0.386) | -0.132 (<0.001) | -0.081 (<0.001) | -0.110 (<0.001) | -0.088 (<0.001) | 0.027 (<0.001) | -0.020 (0.002) | -0.016 (0.014) |
| **Junctional Exponent Deviation** | 0.044 (<0.001) | -0.024 (<0.001) | 0.050 (<0.001) | 0.033 (<0.001) | 0.052 (<0.001) | 0.041 (<0.001) | -0.001 (0.901) | -0.002 (0.731) | -0.000 (0.951) |
| **Branching Coefficient** | -0.039 (<0.001) | 0.010 (0.104) | -0.027 (<0.001) | -0.036 (<0.001) | -0.041 (<0.001) | -0.037 (<0.001) | -0.006 (0.372) | -0.010 (0.114) | 0.008 (0.200) |
| **Branching Angle Measure** | 0.035 (<0.001) | 0.028 (<0.001) | 0.031 (<0.001) | 0.014 (0.025) | 0.016 (0.014) | 0.014 (0.027) | -0.011 (0.087) | 0.008 (0.203) | -0.008 (0.192) |
| **Asymmetry Ratio** | 0.053 (<0.001) | 0.025 (<0.001) | 0.043 (<0.001) | 0.028 (<0.001) | 0.041 (<0.001) | 0.027 (<0.001) | -0.003 (0.601) | 0.006 (0.365) | 0.007 (0.292) |
| **Angular Asymmetry** | 0.023 (<0.001) | 0.039 (<0.001) | 0.036 (<0.001) | -0.022 (<0.001) | 0.027 (<0.001) | -0.011 (0.078) | -0.010 (0.135) | -0.019 (0.003) | 0.024 (<0.001) |

Table 5C. Correlations between retinal vascular features and retinal layer thickness in participants aged 60 and above

|  | Macular thickness  Coefficient (p) | RNFL  Coefficient (p) | GCIPL  Coefficient (p) | INL-RPE  Coefficient (p) | INL  Coefficient (p) | ELM-INL  Coefficient (p) | ISOS-ELM  Coefficient (p) | RPE-ISOS  Coefficient (p) | RPE  Coefficient (p) |
| --- | --- | --- | --- | --- | --- | --- | --- | --- | --- |
| **Tortuosity Density** | 0.039 (<0.001) | 0.007 (0.31) | 0.055 (<0.001) | 0.012 (0.084) | 0.038 (<0.001) | 0.017 (0.011) | -0.029 (<0.001) | 0 (0.949) | 0.011 (0.114) |
| **Inflection Count Tortuosity** | 0.046 (<0.001) | 0.002 (0.733) | 0.071 (<0.001) | 0.012 (0.069) | 0.049 (<0.001) | 0.018 (0.007) | -0.038 (<0.001) | 0.002 (0.771) | 0.012 (0.079) |
| **Fractal Tortuosity** | -0.022 (0.001) | -0.009 (0.205) | -0.031 (<0.001) | 0.001 (0.856) | -0.016 (0.019) | 0.005 (0.491) | 0.021 (0.002) | -0.005 (0.449) | -0.002 (0.758) |
| **Curve Angle** | 0.055 (<0.001) | 0.048 (<0.001) | 0.043 (<0.001) | 0.026 (<0.001) | 0.01 (0.153) | 0.018 (0.007) | -0.003 (0.675) | 0.017 (0.011) | 0.005 (0.44) |
| **Vessel Skeleton Density (vein)** | 0.077 (<0.001) | 0.049 (<0.001) | 0.098 (<0.001) | 0.013 (0.058) | 0.044 (<0.001) | 0.001 (0.916) | -0.024 (<0.001) | 0.02 (0.004) | 0.004 (0.525) |
| **Vessel Skeleton Density (artery)** | 0.102 (<0.001) | 0.047 (<0.001) | 0.118 (<0.001) | 0.029 (<0.001) | 0.073 (<0.001) | 0.012 (0.076) | -0.027 (<0.001) | 0.042 (<0.001) | 0.002 (0.763) |
| **Vessel Area Density (vein)** | 0.086 (<0.001) | 0.034 (<0.001) | 0.114 (<0.001) | 0.021 (0.002) | 0.062 (<0.001) | 0.013 (0.061) | -0.035 (<0.001) | 0.019 (0.006) | 0.004 (0.584) |
| **Vessel Area Density (artery)** | 0.129 (<0.001) | 0.038 (<0.001) | 0.148 (<0.001) | 0.053 (<0.001) | 0.101 (<0.001) | 0.038 (<0.001) | -0.024 (<0.001) | 0.042 (<0.001) | 0.002 (0.75) |
| **Branching Density** | 0.059 (<0.001) | -0.001 (0.868) | 0.059 (<0.001) | 0.044 (<0.001) | 0.037 (<0.001) | 0.04 (<0.001) | -0.022 (<0.001) | 0.021 (0.003) | 0.013 (0.048) |
| **Bifurcation Density** | 0.057 (<0.001) | 0.01 (0.157) | 0.055 (<0.001) | 0.032 (<0.001) | 0.048 (<0.001) | 0.027 (<0.001) | -0.019 (0.006) | 0.022 (0.001) | 0.006 (0.372) |
| **Strahler** | 0.043 (<0.001) | 0.002 (0.824) | 0.068 (<0.001) | 0.011 (0.112) | 0.044 (<0.001) | 0.003 (0.654) | -0.016 (0.02) | 0.02 (0.003) | -0.002 (0.794) |
| **Number of Trees** | -0.015 (0.029) | -0.006 (0.358) | -0.009 (0.172) | -0.011 (0.095) | -0.028 (<0.001) | -0.002 (0.774) | 0.014 (0.041) | -0.013 (0.05) | -0.005 (0.452) |
| **Number of Segments** | 0.065 (<0.001) | -0.034 (<0.001) | 0.133 (<0.001) | 0.016 (0.018) | 0.051 (<0.001) | -0.002 (0.788) | -0.057 (<0.001) | 0.038 (<0.001) | 0.011 (0.107) |
| **Number of Branching** | 0.064 (<0.001) | -0.031 (<0.001) | 0.123 (<0.001) | 0.022 (0.001) | 0.051 (<0.001) | 0.005 (0.476) | -0.05 (<0.001) | 0.038 (<0.001) | 0.007 (0.33) |
| **Number of Bifurcation** | 0.038 (<0.001) | -0.029 (<0.001) | 0.091 (<0.001) | 0.004 (0.51) | 0.025 (<0.001) | -0.01 (0.153) | -0.045 (<0.001) | 0.024 (<0.001) | 0.007 (0.328) |
| **Level** | 0.047 (<0.001) | 0.004 (0.606) | 0.071 (<0.001) | 0.015 (0.026) | 0.046 (<0.001) | 0.01 (0.158) | -0.007 (0.324) | 0.015 (0.023) | 0.005 (0.459) |
| **Width** | 0.106 (<0.001) | -0.032 (<0.001) | 0.128 (<0.001) | 0.082 (<0.001) | 0.115 (<0.001) | 0.087 (<0.001) | -0.01 (0.153) | 0.011 (0.106) | 0.008 (0.227) |
| **Length Diameter Ratio** | -0.113 (<0.001) | 0.007 (0.326) | -0.123 (<0.001) | -0.077 (<0.001) | -0.095 (<0.001) | -0.071 (<0.001) | 0.029 (<0.001) | -0.031 (<0.001) | -0.01 (0.155) |
| **Junctional Exponent Deviation** | 0.045 (<0.001) | -0.014 (0.037) | 0.058 (<0.001) | 0.026 (<0.001) | 0.041 (<0.001) | 0.036 (<0.001) | -0.001 (0.848) | -0.02 (0.003) | 0.021 (0.002) |
| **Branching Coefficient** | -0.031 (<0.001) | 0.006 (0.357) | -0.038 (<0.001) | -0.02 (0.003) | -0.026 (<0.001) | -0.025 (<0.001) | 0.001 (0.851) | 0.01 (0.161) | -0.011 (0.114) |
| **Branching Angle Measure** | 0.03 (<0.001) | 0.029 (<0.001) | 0.023 (<0.001) | 0.014 (0.043) | 0.005 (0.498) | 0.01 (0.134) | -0.013 (0.058) | 0.016 (0.02) | -0.008 (0.259) |
| **Asymmetry Ratio** | 0.05 (<0.001) | 0.017 (0.011) | 0.049 (<0.001) | 0.034 (<0.001) | 0.043 (<0.001) | 0.037 (<0.001) | -0.004 (0.597) | 0.002 (0.791) | 0.012 (0.085) |
| **Angular Asymmetry** | 0.033 (<0.001) | 0.04 (<0.001) | 0.034 (<0.001) | -0.002 (0.744) | 0.027 (<0.001) | 0.006 (0.396) | 0.001 (0.853) | -0.006 (0.379) | 0.001 (0.883) |

Supplementary Table 6. Correlations between retinal vascular features and retinal layer thickness by eye laterality

Table 6A. Correlations between retinal vascular features and retinal layer thickness in right eyes

|  | Macular thickness  Coefficient (p) | RNFL  Coefficient (p) | GCIPL  Coefficient (p) | INL-RPE  Coefficient (p) | INL  Coefficient (p) | ELM-INL  Coefficient (p) | ISOS-ELM  Coefficient (p) | RPE-ISOS  Coefficient (p) | RPE  Coefficient (p) |
| --- | --- | --- | --- | --- | --- | --- | --- | --- | --- |
| **Tortuosity Density** | 0.037 (<0.001) | 0.024 (<0.001) | 0.051 (<0.001) | -0.001 (0.893) | 0.041 (<0.001) | 0.01 (0.053) | -0.032 (<0.001) | -0.01 (0.056) | 0.003 (0.584) |
| **Inflection Count Tortuosity** | 0.047 (<0.001) | 0.02 (<0.001) | 0.074 (<0.001) | 0.001 (0.842) | 0.056 (<0.001) | 0.011 (0.035) | -0.027 (<0.001) | -0.008 (0.119) | 0.012 (0.027) |
| **Fractal Tortuosity** | -0.022 (<0.001) | -0.006 (0.255) | -0.038 (<0.001) | 0.001 (0.885) | -0.026 (<0.001) | -0.004 (0.408) | -0.002 (0.762) | 0.009 (0.089) | -0.02 (<0.001) |
| **Curve Angle** | 0.052 (<0.001) | 0.043 (<0.001) | 0.042 (<0.001) | 0.024 (<0.001) | 0.017 (0.001) | 0.022 (<0.001) | -0.001 (0.869) | 0.009 (0.083) | -0.003 (0.569) |
| **Vessel Skeleton Density (vein)** | 0.092 (<0.001) | 0.044 (<0.001) | 0.139 (<0.001) | 0.01 (0.065) | 0.074 (<0.001) | 0 (0.963) | 0.029 (<0.001) | 0.008 (0.152) | 0.036 (<0.001) |
| **Vessel Skeleton Density (artery)** | 0.123 (<0.001) | 0.047 (<0.001) | 0.173 (<0.001) | 0.024 (<0.001) | 0.106 (<0.001) | 0.007 (0.179) | 0.052 (<0.001) | 0.021 (<0.001) | 0.033 (<0.001) |
| **Vessel Area Density (vein)** | 0.107 (<0.001) | 0.018 (0.001) | 0.162 (<0.001) | 0.027 (<0.001) | 0.099 (<0.001) | 0.022 (<0.001) | 0.014 (0.011) | 0.012 (0.028) | 0.026 (<0.001) |
| **Vessel Area Density (artery)** | 0.154 (<0.001) | 0.04 (<0.001) | 0.205 (<0.001) | 0.051 (<0.001) | 0.133 (<0.001) | 0.036 (<0.001) | 0.051 (<0.001) | 0.028 (<0.001) | 0.028 (<0.001) |
| **Branching Density** | 0.053 (<0.001) | -0.008 (0.127) | 0.078 (<0.001) | 0.028 (<0.001) | 0.048 (<0.001) | 0.032 (<0.001) | 0.01 (0.07) | -0.003 (0.627) | 0.033 (<0.001) |
| **Bifurcation Density** | 0.058 (<0.001) | 0.024 (<0.001) | 0.072 (<0.001) | 0.017 (0.002) | 0.051 (<0.001) | 0.019 (<0.001) | 0.007 (0.228) | -0.001 (0.815) | 0.026 (<0.001) |
| **Strahler** | 0.044 (<0.001) | 0.015 (0.006) | 0.085 (<0.001) | -0.009 (0.104) | 0.049 (<0.001) | -0.003 (0.551) | -0.003 (0.593) | -0.01 (0.074) | 0.019 (<0.001) |
| **Number of Trees** | 0.015 (0.007) | 0.019 (<0.001) | 0.011 (0.047) | 0.003 (0.598) | -0.004 (0.424) | 0.008 (0.149) | 0.023 (<0.001) | -0.009 (0.099) | 0.003 (0.636) |
| **Number of Segments** | 0.061 (<0.001) | -0.058 (<0.001) | 0.175 (<0.001) | -0.007 (0.225) | 0.077 (<0.001) | -0.012 (0.029) | 0.003 (0.527) | -0.004 (0.472) | 0.051 (<0.001) |
| **Number of Branching** | 0.055 (<0.001) | -0.054 (<0.001) | 0.155 (<0.001) | -0.002 (0.696) | 0.07 (<0.001) | -0.01 (0.059) | 0.004 (0.436) | 0.002 (0.767) | 0.045 (<0.001) |
| **Number of Bifurcation** | 0.043 (<0.001) | -0.049 (<0.001) | 0.127 (<0.001) | -0.003 (0.562) | 0.053 (<0.001) | -0.009 (0.08) | 0.003 (0.519) | 0 (0.95) | 0.033 (<0.001) |
| **Level** | 0.046 (<0.001) | 0.017 (0.002) | 0.08 (<0.001) | -0.002 (0.767) | 0.043 (<0.001) | 0 (0.986) | 0.012 (0.03) | -0.004 (0.427) | 0.02 (<0.001) |
| **Width** | 0.111 (<0.001) | -0.042 (<0.001) | 0.141 (<0.001) | 0.08 (<0.001) | 0.119 (<0.001) | 0.088 (<0.001) | -0.011 (0.04) | 0.016 (0.004) | -0.002 (0.765) |
| **Length Diameter Ratio** | -0.116 (<0.001) | 0.008 (0.165) | -0.152 (<0.001) | -0.062 (<0.001) | -0.111 (<0.001) | -0.066 (<0.001) | -0.001 (0.867) | -0.008 (0.159) | -0.032 (<0.001) |
| **Junctional Exponent Deviation** | 0.052 (<0.001) | -0.03 (<0.001) | 0.061 (<0.001) | 0.045 (<0.001) | 0.053 (<0.001) | 0.05 (<0.001) | 0.004 (0.418) | 0.006 (0.29) | 0.009 (0.112) |
| **Branching Coefficient** | -0.039 (<0.001) | 0.03 (<0.001) | -0.044 (<0.001) | -0.043 (<0.001) | -0.042 (<0.001) | -0.04 (<0.001) | -0.007 (0.217) | -0.016 (0.003) | 0.003 (0.538) |
| **Branching Angle Measure** | 0.013 (0.013) | -0.006 (0.298) | 0.015 (0.006) | 0.011 (0.041) | 0.006 (0.256) | 0.011 (0.043) | -0.015 (0.006) | 0.007 (0.218) | -0.01 (0.072) |
| **Asymmetry Ratio** | 0.024 (<0.001) | -0.025 (<0.001) | 0.04 (<0.001) | 0.029 (<0.001) | 0.031 (<0.001) | 0.031 (<0.001) | -0.007 (0.22) | 0.004 (0.465) | 0.011 (0.034) |
| **Angular Asymmetry** | 0.013 (0.014) | 0.022 (<0.001) | 0.036 (<0.001) | -0.02 (<0.001) | 0.02 (<0.001) | -0.014 (0.009) | 0.01 (0.064) | -0.021 (<0.001) | 0.014 (0.009) |

Table 6B. Correlations between retinal vascular features and retinal layer thickness in left eyes

|  | Macular thickness  Coefficient (p) | RNFL  Coefficient (p) | GCIPL  Coefficient (p) | INL-RPE  Coefficient (p) | INL  Coefficient (p) | ELM-INL  Coefficient (p) | ISOS-ELM  Coefficient (p) | RPE-ISOS  Coefficient (p) | RPE  Coefficient (p) |
| --- | --- | --- | --- | --- | --- | --- | --- | --- | --- |
| **Tortuosity Density** | 0.041 (<0.001) | 0.015 (0.006) | 0.055 (<0.001) | 0.002 (0.688) | 0.045 (<0.001) | 0.019 (<0.001) | -0.033 (<0.001) | -0.016 (0.003) | 0.015 (0.006) |
| **Inflection Count Tortuosity** | 0.056 (<0.001) | 0.016 (0.004) | 0.077 (<0.001) | 0.007 (0.202) | 0.058 (<0.001) | 0.021 (<0.001) | -0.028 (<0.001) | -0.011 (0.049) | 0.022 (<0.001) |
| **Fractal Tortuosity** | -0.036 (<0.001) | -0.001 (0.897) | -0.044 (<0.001) | -0.016 (0.004) | -0.03 (<0.001) | -0.01 (0.067) | -0.009 (0.093) | -0.006 (0.294) | -0.021 (<0.001) |
| **Curve Angle** | 0.051 (<0.001) | 0.04 (<0.001) | 0.037 (<0.001) | 0.03 (<0.001) | 0.01 (0.07) | 0.032 (<0.001) | 0.002 (0.701) | 0.007 (0.191) | 0.008 (0.154) |
| **Vessel Skeleton Density (vein)** | 0.101 (<0.001) | 0.054 (<0.001) | 0.128 (<0.001) | 0.021 (<0.001) | 0.07 (<0.001) | 0.004 (0.441) | 0.049 (<0.001) | 0.011 (0.048) | 0.04 (<0.001) |
| **Vessel Skeleton Density (artery)** | 0.125 (<0.001) | 0.039 (<0.001) | 0.165 (<0.001) | 0.033 (<0.001) | 0.096 (<0.001) | 0.012 (0.025) | 0.062 (<0.001) | 0.023 (<0.001) | 0.035 (<0.001) |
| **Vessel Area Density (vein)** | 0.113 (<0.001) | 0.038 (<0.001) | 0.14 (<0.001) | 0.036 (<0.001) | 0.091 (<0.001) | 0.025 (<0.001) | 0.035 (<0.001) | 0.011 (0.039) | 0.033 (<0.001) |
| **Vessel Area Density (artery)** | 0.154 (<0.001) | 0.038 (<0.001) | 0.19 (<0.001) | 0.057 (<0.001) | 0.122 (<0.001) | 0.041 (<0.001) | 0.058 (<0.001) | 0.024 (<0.001) | 0.032 (<0.001) |
| **Branching Density** | 0.07 (<0.001) | -0.01 (0.055) | 0.085 (<0.001) | 0.044 (<0.001) | 0.05 (<0.001) | 0.046 (<0.001) | 0.018 (0.001) | 0.005 (0.324) | 0.035 (<0.001) |
| **Bifurcation Density** | 0.063 (<0.001) | 0.011 (0.037) | 0.069 (<0.001) | 0.029 (<0.001) | 0.051 (<0.001) | 0.032 (<0.001) | 0.01 (0.057) | 0.003 (0.569) | 0.024 (<0.001) |
| **Strahler** | 0.06 (<0.001) | 0.009 (0.08) | 0.089 (<0.001) | 0.012 (0.022) | 0.056 (<0.001) | 0.008 (0.153) | 0.001 (0.804) | 0.012 (0.023) | 0.013 (0.017) |
| **Number of Trees** | 0.001 (0.824) | -0.005 (0.394) | 0.009 (0.099) | -0.003 (0.605) | -0.007 (0.194) | 0.006 (0.269) | 0.015 (0.006) | -0.01 (0.065) | 0.003 (0.638) |
| **Number of Segments** | 0.081 (<0.001) | -0.051 (<0.001) | 0.172 (<0.001) | 0.013 (0.021) | 0.076 (<0.001) | 0.003 (0.612) | 0.027 (<0.001) | 0.007 (0.22) | 0.059 (<0.001) |
| **Number of Branching** | 0.078 (<0.001) | -0.043 (<0.001) | 0.157 (<0.001) | 0.017 (0.002) | 0.069 (<0.001) | 0.007 (0.179) | 0.026 (<0.001) | 0.009 (0.102) | 0.049 (<0.001) |
| **Number of Bifurcation** | 0.046 (<0.001) | -0.045 (<0.001) | 0.119 (<0.001) | 0.001 (0.792) | 0.047 (<0.001) | -0.009 (0.106) | 0.017 (0.002) | 0.004 (0.468) | 0.045 (<0.001) |
| **Level** | 0.07 (<0.001) | 0.017 (0.002) | 0.09 (<0.001) | 0.021 (<0.001) | 0.063 (<0.001) | 0.017 (0.002) | 0.008 (0.14) | 0.011 (0.039) | 0.012 (0.028) |
| **Width** | 0.122 (<0.001) | -0.013 (0.014) | 0.12 (<0.001) | 0.089 (<0.001) | 0.125 (<0.001) | 0.099 (<0.001) | -0.012 (0.025) | 0.015 (0.005) | -0.006 (0.291) |
| **Length Diameter Ratio** | -0.119 (<0.001) | 0.008 (0.128) | -0.134 (<0.001) | -0.074 (<0.001) | -0.101 (<0.001) | -0.079 (<0.001) | -0.009 (0.116) | -0.011 (0.051) | -0.036 (<0.001) |
| **Junctional Exponent Deviation** | 0.042 (<0.001) | -0.012 (0.033) | 0.055 (<0.001) | 0.023 (<0.001) | 0.047 (<0.001) | 0.036 (<0.001) | 0.005 (0.403) | -0.018 (<0.001) | 0.011 (0.036) |
| **Branching Coefficient** | -0.038 (<0.001) | -0.003 (0.622) | -0.035 (<0.001) | -0.027 (<0.001) | -0.033 (<0.001) | -0.028 (<0.001) | -0.013 (0.015) | -0.001 (0.921) | -0.001 (0.889) |
| **Branching Angle Measure** | 0.01 (0.061) | -0.013 (0.02) | 0.01 (0.07) | 0.014 (0.009) | 0.007 (0.199) | 0.022 (<0.001) | -0.025 (<0.001) | 0.004 (0.478) | -0.01 (0.08) |
| **Asymmetry Ratio** | 0.039 (<0.001) | -0.007 (0.18) | 0.035 (<0.001) | 0.031 (<0.001) | 0.04 (<0.001) | 0.034 (<0.001) | -0.003 (0.529) | 0.003 (0.611) | 0.005 (0.354) |
| **Angular Asymmetry** | 0.021 (<0.001) | 0.008 (0.158) | 0.036 (<0.001) | -0.008 (0.156) | 0.029 (<0.001) | 0 (0.998) | 0.003 (0.571) | -0.01 (0.071) | 0.012 (0.024) |

Supplementary Figure 1 Canonical correlations of the first four canonical dimensions

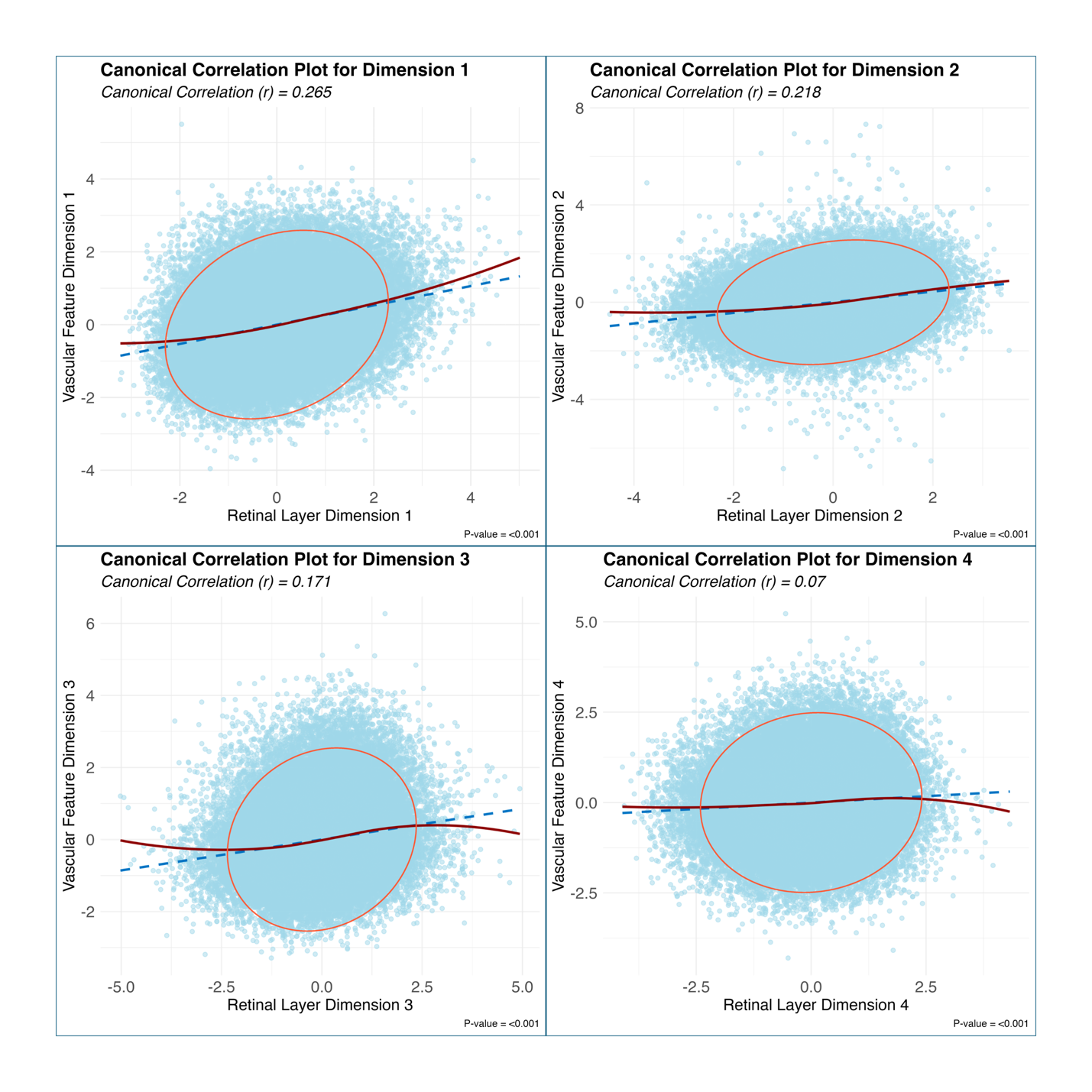

Notes: The red ellipse outlines the 95% confidence region of the data distribution. The blue dashed line shows the linear regression fit, while the solid red curve represents the locally estimated scatterplot smoothing (LOESS) fit, providing a non-linear visualization of the relationship.

Supplementary Figure 2 Loadings of variables in the first four canonical dimensions

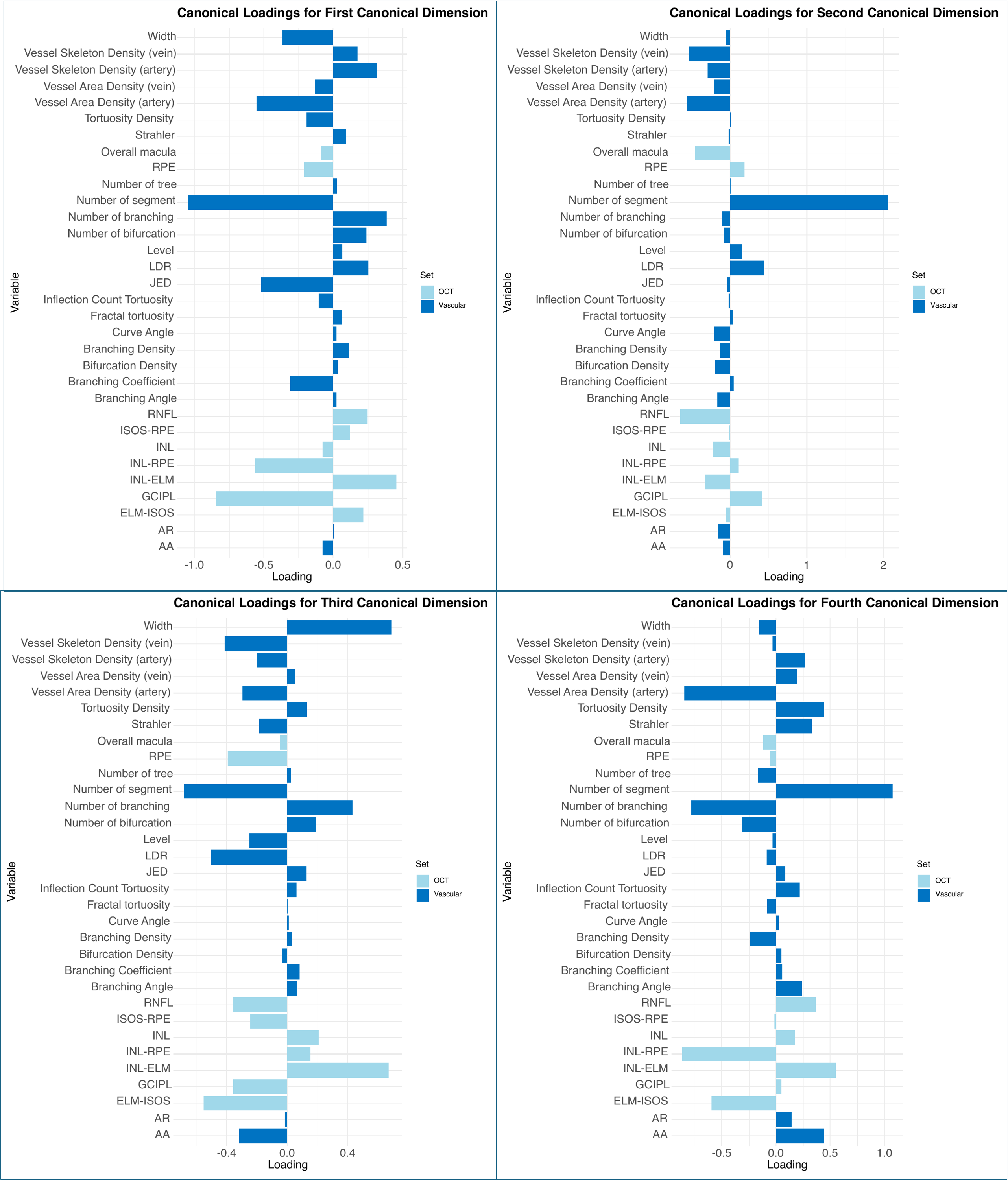

Notes: RNFL = Retinal Nerve Fiber Layer, GCIPL = Ganglion Cell-Inner Plexiform Layer, INL = Inner Nuclear Layer, ELM = External Limiting Membrane, ISOS = Inner Segment/Outer Segment junction, RPE = Retinal Pigment Epithelium. LDR, Length Diameter Ratio; JED, Junctional Exponent Deviation; AA, Angular Asymmetry; AR, Asymmetry Ratio.

Supplementary Figure 3 Scatter plots and forest plots for Mendelian Randomization analysis

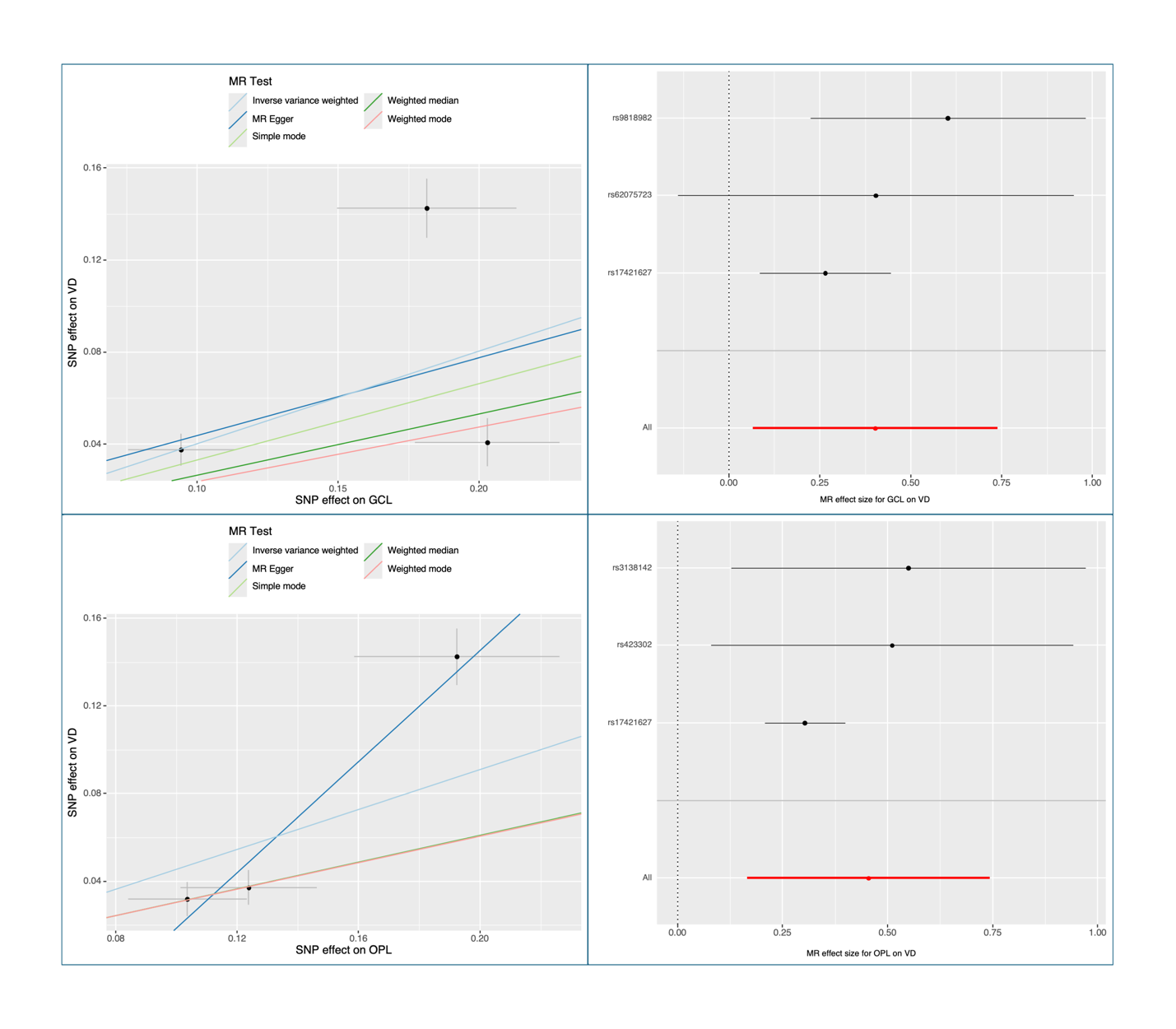

Notes: SNP, single nucleotide polymorphism; VD, Vessel Density; OPL, outer plexiform layer; GCL, ganglion cell layer; MR, Mendelian Randomization.

Supplementary Table 7. Correlations between retinal vascular features and retinal layer thickness as categorical variables

|  | Macular thickness  Coefficient (p) | RNFL  Coefficient (p) | GCIPL  Coefficient (p) | INL-RPE  Coefficient (p) | INL  Coefficient (p) | ELM-INL  Coefficient (p) | ISOS-ELM  Coefficient (p) | RPE-ISOS  Coefficient (p) | RPE  Coefficient (p) |
| --- | --- | --- | --- | --- | --- | --- | --- | --- | --- |
| **Tortuosity Density** | 0.043 (<0.001) | 0.019 (<0.001) | 0.056 (<0.001) | 0.003 (0.32) | 0.046 (<0.001) | 0.017 (0.014) | -0.031 (<0.001) | -0.017 (0.026) | 0.008 (0.643) |
| **Inflection Count Tortuosity** | 0.055 (<0.001) | 0.018 (0.001) | 0.078 (<0.001) | 0.004 (0.006) | 0.059 (<0.001) | 0.016 (<0.001) | -0.029 (<0.001) | -0.013 (<0.001) | 0.019 (<0.001) |
| **Fractal Tortuosity** | -0.028 (<0.001) | -0.003 (0.057) | -0.043 (<0.001) | -0.006 (0.424) | -0.031 (<0.001) | -0.007 (0.234) | -0.005 (0.248) | 0.004 (0.004) | -0.022 (<0.001) |
| **Curve Angle** | 0.062 (<0.001) | 0.062 (<0.001) | 0.042 (<0.001) | 0.026 (<0.001) | 0.017 (<0.001) | 0.026 (<0.001) | 0 (0.055) | 0.009 (0.21) | 0.002 (0.614) |
| **Vessel Skeleton Density (vein)** | 0.11 (<0.001) | 0.07 (<0.001) | 0.146 (<0.001) | 0.015 (0.003) | 0.079 (<0.001) | 0 (<0.001) | 0.044 (<0.001) | 0.01 (<0.001) | 0.039 (<0.001) |
| **Vessel Skeleton Density (artery)** | 0.14 (<0.001) | 0.069 (<0.001) | 0.181 (<0.001) | 0.027 (<0.001) | 0.11 (<0.001) | 0.008 (<0.001) | 0.061 (<0.001) | 0.023 (<0.001) | 0.037 (<0.001) |
| **Vessel Area Density (vein)** | 0.125 (<0.001) | 0.049 (<0.001) | 0.163 (<0.001) | 0.033 (<0.001) | 0.102 (<0.001) | 0.023 (<0.001) | 0.028 (<0.001) | 0.011 (<0.001) | 0.029 (<0.001) |
| **Vessel Area Density (artery)** | 0.173 (<0.001) | 0.064 (<0.001) | 0.213 (<0.001) | 0.055 (<0.001) | 0.139 (<0.001) | 0.039 (<0.001) | 0.06 (<0.001) | 0.026 (<0.001) | 0.033 (<0.001) |
| **Branching Density** | 0.066 (<0.001) | -0.001 (<0.001) | 0.086 (<0.001) | 0.038 (<0.001) | 0.053 (<0.001) | 0.039 (<0.001) | 0.016 (<0.001) | -0.001 (<0.001) | 0.04 (<0.001) |
| **Bifurcation Density** | 0.065 (<0.001) | 0.023 (<0.001) | 0.074 (<0.001) | 0.024 (<0.001) | 0.056 (<0.001) | 0.028 (<0.001) | 0.007 (<0.001) | -0.001 (<0.001) | 0.027 (<0.001) |
| **Strahler** | 0.053 (<0.001) | 0.011 (0.004) | 0.093 (<0.001) | -0.001 (0.019) | 0.055 (<0.001) | 0 (0.219) | 0.002 (0.12) | -0.001 (<0.001) | 0.019 (<0.001) |
| **Number of Trees** | 0.003 (0.065) | -0.005 (0.554) | 0.01 (0.494) | 0 (0.285) | -0.005 (0.625) | 0.006 (0.449) | 0.017 (0.049) | -0.012 (0.127) | 0.004 (0.036) |
| **Number of Segments** | 0.085 (<0.001) | -0.035 (<0.001) | 0.189 (<0.001) | 0.002 (<0.001) | 0.085 (<0.001) | -0.007 (<0.001) | 0.017 (<0.001) | 0 (<0.001) | 0.059 (<0.001) |
| **Number of Branching** | 0.079 (<0.001) | -0.032 (<0.001) | 0.17 (<0.001) | 0.007 (<0.001) | 0.078 (<0.001) | -0.004 (<0.001) | 0.018 (<0.001) | 0.003 (<0.001) | 0.052 (<0.001) |
| **Number of Bifurcation** | 0.054 (<0.001) | -0.032 (<0.001) | 0.132 (<0.001) | -0.003 (0.109) | 0.055 (<0.001) | -0.01 (0.015) | 0.012 (<0.001) | -0.001 (<0.001) | 0.042 (<0.001) |
| **Level** | 0.06 (<0.001) | 0.015 (0.001) | 0.093 (<0.001) | 0.012 (0.203) | 0.057 (<0.001) | 0.01 (0.027) | 0.015 (0.019) | 0.004 (<0.001) | 0.019 (0.003) |
| **Width** | 0.123 (<0.001) | -0.026 (<0.001) | 0.138 (<0.001) | 0.088 (<0.001) | 0.129 (<0.001) | 0.098 (<0.001) | -0.011 (0.001) | 0.014 (<0.001) | -0.001 (0.262) |
| **Length Diameter Ratio** | -0.126 (<0.001) | 0 (<0.001) | -0.152 (<0.001) | -0.071 (<0.001) | -0.111 (<0.001) | -0.075 (<0.001) | -0.008 (<0.001) | -0.009 (<0.001) | -0.038 (<0.001) |
| **Junctional Exponent Deviation** | 0.049 (<0.001) | -0.024 (<0.001) | 0.063 (<0.001) | 0.037 (<0.001) | 0.052 (<0.001) | 0.049 (<0.001) | 0.006 (0.005) | -0.008 (0.064) | 0.009 (0.078) |
| **Branching Coefficient** | -0.041 (<0.001) | 0.01 (<0.001) | -0.043 (<0.001) | -0.036 (<0.001) | -0.037 (<0.001) | -0.036 (<0.001) | -0.012 (0.017) | -0.007 (0.025) | 0.003 (0.664) |
| **Branching Angle Measure** | 0.033 (<0.001) | 0.035 (<0.001) | 0.019 (<0.001) | 0.012 (0.113) | 0.008 (0.059) | 0.014 (0.005) | -0.02 (0.003) | 0.006 (0.051) | -0.009 (0.587) |
| **Asymmetry Ratio** | 0.05 (<0.001) | 0.025 (<0.001) | 0.043 (<0.001) | 0.028 (<0.001) | 0.039 (<0.001) | 0.031 (<0.001) | -0.008 (0.025) | -0.002 (0.78) | 0.011 (0.051) |
| **Angular Asymmetry** | 0.034 (<0.001) | 0.053 (<0.001) | 0.041 (<0.001) | -0.013 (0.026) | 0.028 (<0.001) | -0.01 (0.605) | 0.008 (0.261) | -0.015 (0.012) | 0.015 (0.002) |
